# Plitidepsin has a positive therapeutic index in adult patients with COVID-19 requiring hospitalization

**DOI:** 10.1101/2021.05.25.21257505

**Authors:** José F. Varona, Pedro Landete, Jose A. Lopez-Martin, Vicente Estrada, Roger Paredes, Pablo Guisado-Vasco, Lucía Fernández de Orueta, Miguel Torralba, Jesús Fortún, Roberto Vates, José Barberán, Bonaventura Clotet, Julio Ancochea, Daniel Carnevali, Noemí Cabello, Lourdes Porras, Paloma Gijón, Alfonso Monereo, Daniel Abad, Sonia Zúñiga, Isabel Sola, Jordi Rodon, Nuria Izquierdo-Useros, Salvador Fudio, María José Pontes, Beatriz de Rivas, Patricia Girón de Velasco, Belén Sopesén, Antonio Nieto, Javier Gómez, Pablo Avilés, Rubin Lubomirov, Kris M. White, Romel Rosales, Soner Yildiz, Ann-Kathrin Reuschl, Lucy G. Thorne, Clare Jolly, Greg J. Towers, Lorena Zuliani-Alvarez, Mehdi Bouhaddou, Kirsten Obernier, Luis Enjuanes, Jose M. Fernández-Sousa, Plitidepsin – COVID - 19 Study Group, Nevan J. Krogan, José M. Jimeno, Adolfo García-Sastre

## Abstract

Plitidepsin is a marine-derived cyclic-peptide that inhibits SARS-CoV-2 replication at low nanomolar concentrations by the targeting of host protein eEF1A (eukaryotic translation-elongation-factor-1A). We evaluated a model of intervention with plitidepsin in hospitalized COVID-19 adult patients where three doses were assessed (1.5, 2 and 2.5 mg/day for 3 days, as a 90-minute intravenous infusion) in 45 patients (15 per dose-cohort). Treatment was well tolerated, with only two Grade 3 treatment-related adverse events observed (hypersensitivity and diarrhea). The discharge rates by Days 8 and 15 were 56.8% and 81.8%, respectively, with data sustaining dose-effect. A mean 4.2 log10 viral load reduction was attained by Day 15. Improvement in inflammation markers was also noted in a seemingly dose-dependent manner. These results suggest that plitidepsin impacts the outcome of patients with COVID-19.

**One-Sentence Summary:** Plitidepsin, an inhibitor of SARS-Cov-2 *in vitro*, is safe and positively influences the outcome of patients hospitalized with COVID-19.

## Introduction

As of April 2021, there have been approximately 135 million confirmed cases of COVID-19 reported to the World Health Organization (WHO), including 3 million deaths. *(1)* The lack of effective antiviral therapies represents a glaring unmet need, not only for the treatment of the current severe acute respiratory syndrome coronavirus 2 (SARS-CoV-2) pandemic, *(2)* but also for potential future pandemics, which may originate from other emergent coronaviruses. *(3-5)*

Viruses, particularly single-stranded positive-sense RNA viruses, are ubiquitous in the sea, where they participate directly or indirectly in the population dynamics of marine organisms. *(6)* Thus, the sea may prove to be a reservoir of potent, naturally selected antiviral compounds.

One such compound, plitidepsin, is a cyclic depsipeptide originally isolated from a Mediterranean marine tunicate (*Aplidium albicans*) and is structurally related to didemnins, some of which (i.e., those isolated from *Trididemnum solidum*) have shown antiviral properties. *(7, 8)*

Plitidepsin targets eukaryotic translation elongation factor 1A (eEF1A) *(9)*, which is one of the most abundant protein synthesis factors in the eukaryotic cell, *(9, 10)* and a protein known to be used by viruses to facilitate their replication inside their host. *(11)* The SARS-CoV-2 nucleocapsid (N) protein is a key element involved in packaging of the viral RNA genome, *(12, 13)* and interacts with eEF1A. This interaction might be essential for viral replication, as eEF1A knockdown results in a significant reduction in virus replication. *(12, 13)*

Originally developed as a cancer treatment, plitidepsin has undergone an extensive clinical development program. Specifically, several phase I and II clinical trials have been conducted to explore different intravenous dosing schedules and infusion times *(14-18)*. Pharmacokinetic and safety properties of plitidepsin were gathered from these studies. Based on the results obtained from a phase III clinical trial (ADMYRE) *(19)*, the Australian Therapeutic Goods Administration (TGA) approved the combination of plitidepsin with dexamethasone for the treatment of patients with relapsed/refractory multiple myeloma in 2018. *(20)*

In this study, we describe the results from a Proof-of Concept clinical trial that explores the potential of plitidepsin as a therapy for patients hospitalized with COVID-19.

## Results

We have evaluated the antiviral activity of plitidepsin against different coronavirus species, strains and variants (Materials and Methods). Treatment of Huh-7 cells with as little as 0.5nM of plitidepsin inhibited infection of a human coronavirus 229E expressing GFP (Figure 1A). A 10^4^-fold decrease in SARS-CoV genomic RNA (gRNA) accumulation and a 10^3^-fold decrease in virus SARS-CoV titers were observed in VeroE6 cells treated with 50nM plitidepsin. (Figure S1 and Table S1). By comparison, and consistent with previous results *(10, 21)* plitidepsin showed nanomolar efficacy against SARS-CoV-2-induced cytopathic effects on Vero E6 cells with a half-maximal inhibitory concentration (IC_50_) of 0.038µM, at concentrations where no cytotoxic effects were observed (CC_50_ 2.9µM) (Figure 1B). Moreover, plitidpesin maintained its nanomolar potency against replication of early as well as recently emerging SARS-CoV-2 lineages, such as B.1.1.7, in human lung and gastrointestinal cell lines (Figure 1C). Noteworthy, plitidepsin was more effective against both variants than remdesivir (Figure 1C).

**Figure 1.**
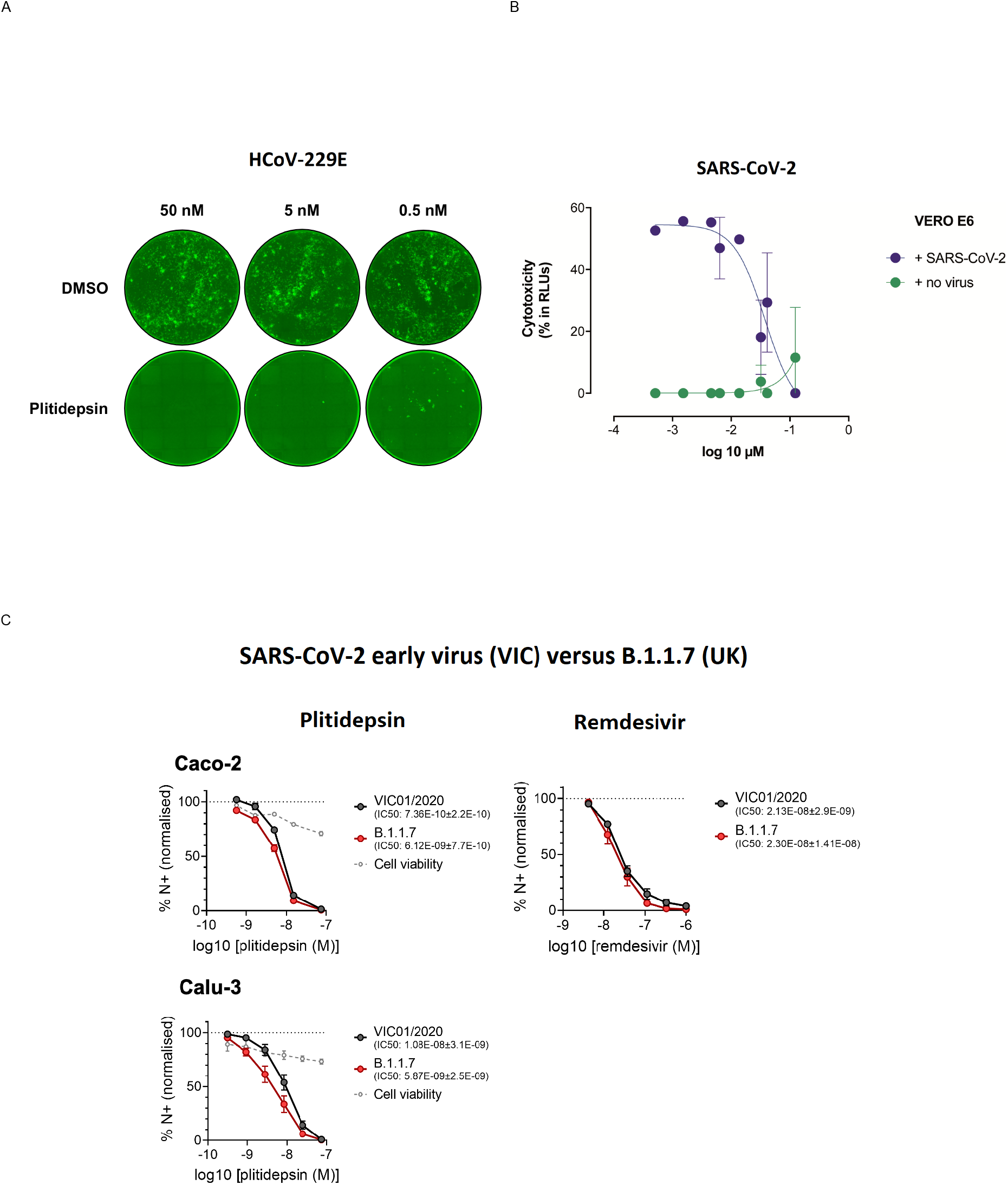
Plitidepsin shows strong antiviral activity *in vitro* against different coronavirus species and variants. (A) HCov-229E-GFP-infected Huh-7 cells were treated with indicated doses of plitidepsin or DMSO. All cells were treated 8 hours after infection and fluorescent foci were analyzed at 48 hours. (B) Cytopathic effect on Vero E6 cells exposed to a fixed concentration of SARS-CoV-2 in the presence of increasing concentrations of plitidepsin. Drug was used at a concentration ranging from 5 nM to 100 μM. Non-linear fit to a variable response curve from one representative experiment with two replicates is shown (blue), excluding data from drug concentrations with associated toxicity; cytotoxicity in the absence of virus is also shown (green). (C) Calu-3 and Caco-2 cells were pre-treated with remdesivir or plitidepsin at the indicated concentrations or DMSO control at an equivalent dilution for 2 h before SARS-CoV-2 infection. Cells were harvested after 24h for analysis, and viral infection measured by intracellular detection of SARS-CoV-2 nucleoprotein by flow cytometry. Tetrazolium salt (MTT) assay was performed to verify cell viability.

To identify target human plasma concentrations of plitidepsin for SARS-CoV-2 infection we developed an extrapolation from *in vitro* results, in line with current recommendations. *(22)* This approach integrated results from non-clinical studies described under Material and Methods (Supplement), including *in vitro* drug sensitivity data for SARS-CoV-2 in Vero cells, human plasma protein binding data (98%) (Table S2), and *in vivo* tissue distribution data in rats (lung-to-plasma partition coefficient ratio of 543-fold (Table S3).

The target plasma and lung concentrations for plitidepsin were initially based on *in vitro* data obtained by Boryung Pharmaceuticals, stablishing IC_50_ of 3.26 nM and IC_90_ of 9.38 nM (Material & Methods). A validated pharmacokinetic population model of plitidepsin *(23)* was used to simulate plasma exposures at different dose levels and infusion durations, so that plitidepsin plasma profiles would reach 0.33 µg/L, and 0.96 µg/L, assuring target concentrations in lung above the aforementioned *in vitro* IC_50_ and IC_90_. A 3-day daily schedule was initially selected to achieve sustained active exposures, under the hypothesis that an acute reduction of the viral load would prevent the onset of the more severe inflammatory phase of COVID-19. The predicted plasma concentrations of plitidepsin, at a dose of 1.5 mg infused intravenously over 90 min, were above the target IC_50_ for the full treatment period and above the IC_90_ for half of the treatment period. The respective predictions after a dose of 2.5 mg were above the IC_90_ during most of the treatment period (Figure 2). Thus, we anticipated that the proposed range of doses would result in stable active concentrations in critical anatomical compartments, such as lung, for more than 120 hours. This model was later supported by White et al, who reported an IC_90_ of 0.88 nM *(10)*. To reach this target concentration in lung tissue, according to the above reasoning, plitidepsin plasma concentration should be above 0.18 µg/L.

**Figure 2.**
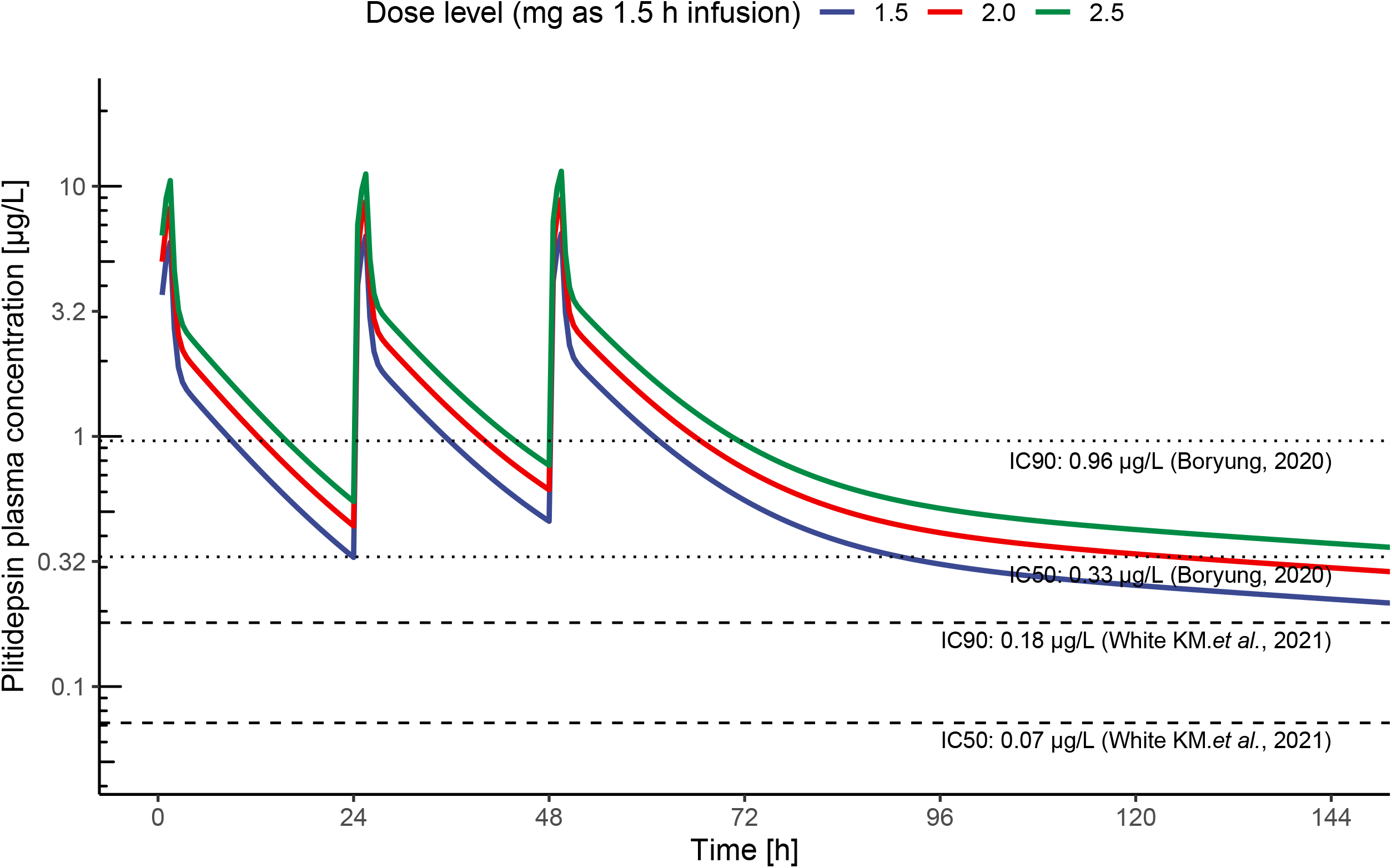
Pharmacological estimation of active plasma concentrations of plitidepsin. Predicted plasma concentrations achieved by a 90 min IV infusion of plitidepsin (1.5, 2 and 2.5 mg) and plasma IC_50_ and IC_90_ thresholds to assure concentrations in lung above IC_50_ and IC_90_ stablished *in vitro*, respectively. *(10, 41)*. Results from Boryung Pharmaceuticals (Material & Methods) were obtained first and used to support the study doses and schedule.

We subsequently designed the APLICOV-PC study [APL-D-002-20; EudraCT #2020-001993-31; NCT #04382066] as a proof-of-concept clinical trial, exploring three dose levels of plitidepsin (1.5 mg/day, 2.0 mg/day, and 2.5 mg/day, flat doses) for 3 consecutive days, as a 90-min intravenous (IV) infusion, in adult patients with COVID-19 who required hospitalization. The primary endpoints were related to safety, but secondary efficacy and pharmacodynamic endpoints were also included (Material and Methods).

Enrollment began on May 12, 2020 and the study was completed by November 26, 2020. Here, we present the data analyses by the cut-off date of December 10, 2020.

In total, 46 hospitalized COVID-19 patients were enrolled across 10 sites in Spain. Baseline demographic and clinical characteristics are summarized in Table 1. One patient withdrew consent before initiating study-specific procedures. The 45 patients who received treatment were sequentially allocated to one of the three dose cohorts; whenever more than 1 dose cohort was open, a patient was assigned by central randomization (Material and Methods).

**Table 1.** Patients’ Baseline Characteristics.

| Parameter | 1.5 mg/day<sup>a</sup> | 2 mg/day<sup>a</sup> | 2.5 mg/day<sup>a</sup> |
| --- | --- | --- | --- |
| Age (years)<sup>b</sup> | 51 (32-75) | 49 (34-71) | 53 (31-84) |
| Gender - N (%) |  |  |  |
| Male | 11 (73.3 %) | 11 (73.3 %) | 8 (53.3 %) |
| Female | 4 (26.7 %) | 4 (26.7 %) | 7 (46.7 %) |
| Ethnic Group - N (%) |  |  |  |
| White | 13 (86.7 %) | 9 (60 %) | 9 (60 %) |
| Latino | 2 (13.3 %) | 4 (26.7 %) | 6 (40 %) |
| Asian | 0 (0 %) | 1 (6.7 %) | 0 (0 %) |
| Arab | 0 (0 %) | 1 (6.7 %) | 0 (0 %) |
| Time from symptom onset to first administration(days)<sup>b</sup> | 6 (3-10) | 6 (3-10) | 6 (2-10) |
| Comorbidities - N (%) |  |  |  |
| One | 2 (13.3 %) | 7 (46.7 %) | 6 (40 %) |
| Two or more | 8 (53.3 %) | 6 (40 %) | 7 (46.7 %) |
| Hypertension | 2 (13.3 %) | 2 (13.3 %) | 5 (33.3 %) |
| Heart disease | 1 (6.7 %) | 0 (0 %) | 1 (6.7 %) |
| COPD<sup>c</sup> | 1 (6.7 %) | 1 (6.7 %) | 1 (6.7 %) |
| Asthma | 2 (13.3 %) | 0 (0 %) | 3 (20 %) |
| Kidney disease | 0 (0 %) | 1 (6.7 %) | 0 (0 %) |
| Diabetes | 1 (6.7 %) | 5 (33.3 %) | 2 (13.3 %) |
| Obesity | 1 (6.7 %) | 5 (33.3 %) | 4 (26.7 %) |
| Disease severity at entry - N (%) (24) |  |  |  |
| Mild COVID-19 | 2 (13.3 %) | 3 (20 %) | 1 (6.7 %) |
| Moderate COVID-19 | 8 (53.3 %) | 7 (46.7 %) | 8 (53.3 %) |
| Severe COVID-19 | 5 (33%) | 5 (33%) | 6 (40%) |
| SpO<sub>2</sub> at room air - N (%) |  |  |  |
| > 93% | 8 (88.9 %) | 6 (85.7 %) | 6 (100 %) |
| ≤ 93% | 1 (11.1 %) | 1 (14.3 %) | 0 (0 %) |
| SpO<sub>2</sub> at room air (%)<sup>b</sup> | 95 (92-99) | 95 (91-97) | 96.5 (94-97) |
| D-dimer (ng/mL)<sup>b</sup> | 330<br>(162-1081) | 463.5<br>(200-1270) | 415<br>(106-962) |
| Ferritin (ng/mL)<sup>b</sup> | 408<br>(96.8-1652.8) | 597<br>(174-1055.2) | 411.9<br>(12.2-1647) |
| C-Reactive Protein (mg/L)<sup>b</sup> | 16.7<br>(1.2-120.4) | 60.7<br>(2.1-128) | 32.7<br>(0.3-120) |
| log<sub>10</sub> copies/ml viral load median (range)<sup>b</sup> | 6.3 (0-9.7) | 6.2 (3.8-7) | 5.7 (0-10.6) |
| Six-point ordinal scale – N (%) (25) |  |  |  |
| 2<sup>d</sup> | 6 | 6 | 4 |
| 3<sup>e</sup> | 8 | 9 | 11 |

<sup>a</sup> Daily x 3 consecutive days<sup>b</sup> Median (range)<sup>c</sup> Chronic Obstructive Pulmonary Disease<sup>d</sup> Hospital admission, not requiring supplemental oxygen<sup>e</sup> Hospital admission, requiring supplemental oxygen

Forty-four patients completed the study through day 31, with one patient in the 1.5 mg cohort withdrawing from the study after the first infusion due to a grade 3 hypersensitivity reaction. The average patient age was 52 years (range 31–84 years). Most patients were male (66.7%) and 80% had co-morbidities (46.7% had two or more). The most commonly reported comorbidities were obesity (22.2%), hypertension (20%), and type 2 diabetes mellitus (17.8%). The distribution of comorbidities was similar between the three treatment cohorts.

Most patients had moderate COVID-19 (51.1%), according to FDA categorization *(24)*, with 13.3% and 35.6% having mild and severe disease, respectively. Baseline chest X-rays showed evidence of lower respiratory infection (infiltrates, unilateral pneumonia, or bilateral pneumonia) in 41 of 45 treated patients (91%), with bilateral pneumonia seen in 32 of them (71%); the percentage of patients with bilateral pneumonia was similar across dose cohorts. Viral load was similar across the three cohorts, with average baseline values for SARS-CoV-2 RNA from nasopharyngeal samples of 6.1 log_10_ copies/mL as measured by quantitative reverse transcriptase polymerase chain reaction (qRT-PCR).

Overall, plitidepsin treatment was well tolerated. As noted above, 1 patient discontinued treatment after a grade 3 hypersensitivity reaction to the first infusion of plitidepsin. The study protocol was thereafter amended to include IV premedication with dexamethasone 8 mg and antihistamine drugs (anti-H1 and anti-H2), 30-min before the infusion of the study drug.

All 45 of the treated patients were evaluable for safety. Nearly all (44 of 45) patients experienced one or more adverse event (AEs). These AEs were determined to be treatment related in only 25 patients (55.5%). Regardless of causality, 14 (31%) patients experienced at least one Grade ≥ 3 AE according to National Cancer Institute – Common Toxicity Criteria for Adverse Events, version 5.0 (NCI-CTCAE v5). The prevalence of grade 3-4 AEs was 40.0% in the 2.5 mg cohort, 20.0% in the 2.0 mg cohort, and 33.3% in the 1.5 mg cohort. Though almost all Grade ≥ 3 AEs were attributed to COVID-19, two Grade 3 AEs were attributed to plitidepsin: one case each of anaphylactic reaction (at 1.5 mg/day) and diarrhea (at 2.5 mg/day). No Grade 4 AEs were reported in patients receiving plitidepsin.

Table 2 presents the frequency of treatment-related AEs in this study. The following treatment-related AEs occurred in more than one patient: nausea (42.2%), vomiting (15.6%), diarrhea (6.7%), abdominal pain (4.4%), dizziness (4.4%), and dysgeusia (4.4%). These events were all mild to moderate (Grade 1-2) except the one case of Grade 3 diarrhea described above. The implementation of the aforementioned protocol amendment was associated with a reduction in the proportion of patients with nausea (from 55.6% to 38.9%) and vomiting (from 22.2% to 13.9%). No new hypersensitivity reactions were seen in any of the 36 patients treated after the amendment (108 infusions).

**Table 2.** Plitidepsin-Related Adverse Events.

| Parameter | Pre-Amendment<sup>a,b</sup> |  |  | Post-Amendment<sup>c</sup> |  |  |
| --- | --- | --- | --- | --- | --- | --- |
|  | Grade 1<br>N (%) | Grade 2<br>N (%) | Grade 3<br>N (%) | Grade 1<br>N (%) | Grade 2<br>N (%) | Grade 3<br>N (%) |
| Nausea | 3 (33.3%) | 2 (22.2%) | - | 11 (30.6%) | 3 (8.3%) | - |
| Vomiting | 2 (22.2%) | - | - | 3 (8.3%) | 2 (5.6%) | - |
| Diarrhea | - | - | - | 1 (2.8%) | 1 (2.8%) | 1 (2.8%) |
| Abdominal Pain | - | - | - | 2 (5.6%) | - | - |
| Dyspepsia | - | - | - | 2 (5.6%) | - | - |
| Asthenia | - | - | - | 1 (2.8%) | 1 (2.8%) | - |
| Anorexia | - | - | - | 1 (2.8%) | - | - |
| Chest Discomfort | - | - | - | 1 (2.8%) | - | - |
| Temperature Regulation Disorder | - | - | - | 1 (2.8%) | - | - |
| Dysthermia | - | - | - | 1 (2.8%) | - | - |
| Anaphylactic Reaction | - | - | 1 (11.1%) | - | - | - |
| Amylase Increase<sup>d</sup> | - | - | - | - | 1 (2.8%) | - |
| Lipase Increase<sup>e</sup> | - | - | - | - | 1 (2.8%) | - |
| Decreased Appetite | - | - | - | - | 1 (2.8%) | - |
| Dizziness | - | - | - | 2 (5.6%) | - | - |
| Dysgeusia | - | - | - | 2 (5.6%) | - | - |

<sup>a</sup> Relevant amendment #9 was implemented in Protocol v5.0 dated 13 August 2020: It modified prophylactic medication prior to plitidepsin infusion to add ondansetron 8 mg IV slow infusion and changed the route of administration of dexamethasone, from oral to IV. The dose of dexamethasone was 8 mg (calculated as 8 mg dexamethasone phosphate, which is equivalent to 6.6 mg dexamethasone base).
<sup>b</sup> 9 patients; 25 plitidepsin IV infusions.
<sup>c</sup> 36 patients; 108 plitidepsin IV infusions.
<sup>d</sup> Short lasting, five minutes, retro sternal low intensity pain during 1st day IV infusion: self-resolved plitidepsin infusion completed day 1, 2 and 3.
<sup>e</sup> Same patient, onset day 2 Plitidepsin Therapy self-resolved in 48 hours.

Several laboratory abnormalities were reported in these patients, most of which were consistent with the acute, inflammatory nature of COVID-19. Of note, plitidepsin did not show signs of clinically relevant hemotoxicity; there were two patients with neutropenia (one Grade 1 and another Grade 2). An isolated observation of Grade 3 neutropenia was reported in an asymptomatic outpatient on the Day 31 follow-up; this patient was also taking metamizole, and the responsible investigator deemed that the event was neither clinically relevant nor related to plitidepsin. Of the 32 patients who entered with normal platelet counts, only one had Grade 1 thrombocytopenia, whereas of the 12 patients who entered into the study with Grade 1 thrombocytopenia, 5 (41.7%) had counts normalized.

Abnormalities in liver function tests were common, transient, mild and reversible. Elevation of alanine aminotransferase (ALT) and aspartate aminotransferase (AST) were reported in 29 of 44 (66%) and 13 of 44 (30%) patients, respectively. Two patients had a single and self-limited observation of a Grade 3 increase in ALT, with no associated increase in bilirubin. Four patients of the 42 who entered with normal creatinine developed a Grade 1 increase in creatinine on study. A Grade 1 increase in creatinine phosphokinase (CPK) was documented in 3 of 37 patients (8.1%) who had normal baseline values, whereas 4 of 5 patients (80%) who entered with Grade 1-2 elevation had their CPK values decreased on study.

Hyperglycemia was documented in 8 of 45 patients (18%): 3 patients in the 1.5 mg/day dose cohort, 2 patients in the 2 mg/day, and 3 patients in the 2.5 mg/day group. All cases of hyperglycemia were Grade ≤ 2 except one patient, who had Grade 3 hyperglycemia lasting for 2 days. For 5 patients, hyperglycemia was considered related to concomitant medication. None of these events were considered to be related to plitidepsin.

Three patients in this study died (6.7%); all had severe disease at baseline, and each death was determined to be related to COVID-19. Deaths occurred on Days 22, 30 and 57 after the start of treatment with plitidepsin. One patient received plitidepsin 1.5 mg/day and the other two received 2.5 mg/day, with no reported tolerability issues.

Seven additional patients experienced serious AEs (SAEs): five dosed at 1.5 mg/day, 1 dosed at 2.0 mg/day, and 1 dosed at 2.5 mg/d. As previously mentioned, only one SAE (2.2% subjects) was considered related to the study drug: a Grade 3 hypersensitivity reaction occurring 5 minutes after the start of the first infusion of plitidepsin. This patient was withdrawn from the study before completing the full treatment.

All 44 patients who completed the 3-day treatment with plitidepsin were evaluated for efficacy analyses. Overall, the discharge rates by Days 8 and 15 after the start of plitidepsin were 56.8% (25 of 44) and 81.8% (36 of 44), respectively. Likewise, a mean -4.2 log_10_ reduction in viral load from baseline was attained by Day 15. Without adjusting for any covariates and with the constraints of the small sample size, there was not a clear dose effect on discharge rates at Day 15: 78.6% (1.5 mg/day), 93.3% (2 mg/day) and 73.3% (2.5 mg/day). Notably, the proportion of patients achieving hospital discharge by Day 8 seemed to increase with dose, from 42.9% for those receiving 1.5 mg/day to 60% and 66.7% for those receiving 2 and 2.5 mg/day, respectively (Table S4).

Figure 3A plots the length of the hospitalization and time in the intensive care unit (ICU) per subject, dose and severity of the disease. As expected, the length of hospitalization was greater in patients with severe disease at baseline. The median time to discharge was 7 days (interquartile range [IQR]: 7-9 days) in patients with mild COVID-19, 7 days (IQR: 6-8 days) with moderate COVID-19, and 14 days (IQR: 7-26 days) with severe COVID-19 at baseline (log-rank p=0.001) (Figure 3B). The discharge rate by Day 15 was 95.7% (22 of 23) in patients with moderate disease compared with 53.3% (8 of 15) in patients with severe disease, and 100% (6 of 6) in patients with mild disease. Similarly, the discharge rate by Day 8 was 73.9% (17 of 23) in patients with moderate disease compared with 27% (4 of 15) in patients with severe disease, and 66.7% (4 of 6) in patients with mild disease.

**Figure 3.**
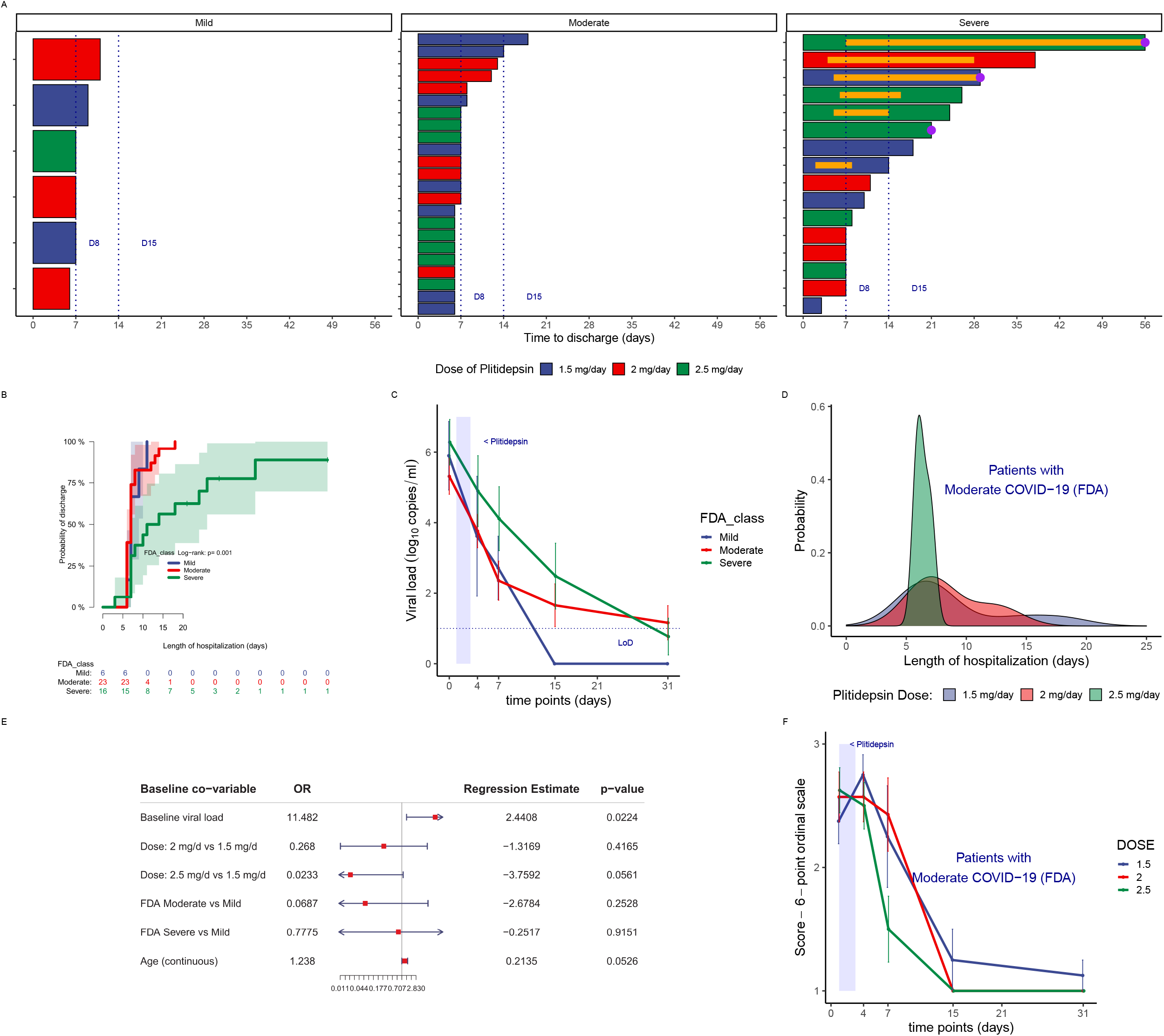
APLICOV-PC Study: Efficacy Outcomes. (A) Length of hospitalization, by disease severity at baseline *(24)* and dose of plitidepsin. Orange bars represent admission in Intensive Care Units. Purple circles represent the three deaths reported in the study. (B) Reverse Kaplan Meier plot showing the cumulative incidence of hospital discharge by baseline severity (mild, moderate or severe COVID-19, according to FDA definition). *(24)* (C) Viral load kinetics (qRT-PCR from nasopharyngeal exudates), by baseline severity.*(24)* (D) Temporal distribution of the probability of discharge in patients with moderate COVID-19 at baseline, according to the dose of plitidepsin administered. (E) Exploratory logistic regression model for hospital discharge at Day 8, with selected covariates. Baseline viral load negatively predicts poorer outcome. (F) Mean score over time of the 6-category ordinal scale *(25)* in patients with moderate disease at baseline, according to the administered dose of plitidepsin. See also Table S5 and Figure S6.

Non-mechanical invasive ventilation was required in one patient with moderate disease (4.3%) and in seven patients with severe disease (46.7%) (p= 0.003). Six patients required intensive care support (13.6%), all of whom had severe disease at baseline (6 of 15, 40%). Tables S4 – S6 summarize additional outcome measures.

Most patients in this study presented with moderate COVID-19 at baseline. All patients in this subgroup (8 of 8) who were allocated to the highest dose level (2.5 mg/day) were discharged by day 8, whereas 3 of 7 patients (43%) and 3 of 8 patients (38%) treated at 2.0 mg/day and 1.5 mg/day, respectively, were discharged beyond that time point. These data are visualized in Figure 3D, which shows that for patients with moderate disease who received plitidepsin 2.5 mg/day, the most probable duration of hospitalization was approximately 1 week with narrow variation with respect to the other doses.

While on study, 64.4% (29 of 45) of patients received systemic corticosteroids, besides its use as pre-medication. After completing plitidepsin treatment, 15.5% (7 of 45) of patients received additional treatments for COVID-19, including the anti-viral agent remdesivir (1 patient) and/or the anti-IL-6 monoclonal antibody tocilizumab (6 patients).

After their treatment, patients’ viral loads were evaluated by RT-PCR. Viral load showed mean declines from baseline of 1.67, 2.84, and 4.24 log_10_ copies/mL at days 4, 7, and 15, respectively (Table S4; see also Table S7 for individual assessments). The mean time to undetectable viral load was 13 days, longer in patients with severe disease at baseline (15 days) respect to mild or moderate disease (12 days) (Figure 3C). In the primary analysis, there were no significant differences in change to viral load across dose levels (Figure S4).

Baseline viral load was found to be significantly correlated with hospital discharge by Day 15, by logistic and Cox regression models. This occurred despite the limitations for modeling (due to the small number of patients [N=44] and the high percentage of patients [82%] discharged by Day 15), following a stepwise selection of covariates with a p-value <0.10 in univariate logistic models. As it looked like Day 8 could be more informative and discriminative for a new drug in COVID-19, as well as more relevant from a clinical point of view, a new *post-hoc* model was built. Twenty-nine patients (64.4%) had a favorable outcome. Given the small sample size, the baseline variables we selected for exploratory purposes were limited to age, viral load, disease severity, and dose cohort. Figure 3E shows the coefficients and Forest Plot for this model. Again, increased baseline viral load significantly correlated with decreased hospital discharge rate by Day 8. Notwithstanding, allocation to the plitidepsin 2.5 mg/day dose cohort and a younger age had higher probabilities of positive outcome, with the constraints of small sample size.

As the cohort of patients with moderate COVID-19 at baseline was the largest, and given that the above model assigned to the variable ‘dose of plitidepsin’ a marginally predictive weight, we focused on that category to capture hints of dose-related activity with which hypotheses could be built for future development of plitidepsin. Although statistical analyses were not performed, due to the small sample size, visual exploration of mean trends do not appear to capture dose-dependent differences in the kinetics of viral PCR, in the moderate COVID-19 subgroup (Figure 4I). Nevertheless, it suggests that the higher the dose of plitidepsin, the smaller the peak of C-reactive protein at day 7 (Figure 4H), the faster the increase in lymphocyte count (Figure 4G), and the faster the recovery of neutrophil-to-lymphocyte ratio (Figure 4J), D-Dimer (Figure 4K), and in the score of a 6-ordinal scale for outcome (Figure 3F and Figure S6).*(25)* These findings may reflect immunomodulatory/anti-inflammatory effects, that could be either secondary to the reduction in viral load or plitidepsin-mediated, and might participate in the rapid recovery of lung infiltrates reported in some patients with computed tomography scan evaluations (Figure 4ABC).

**Figure 4.**
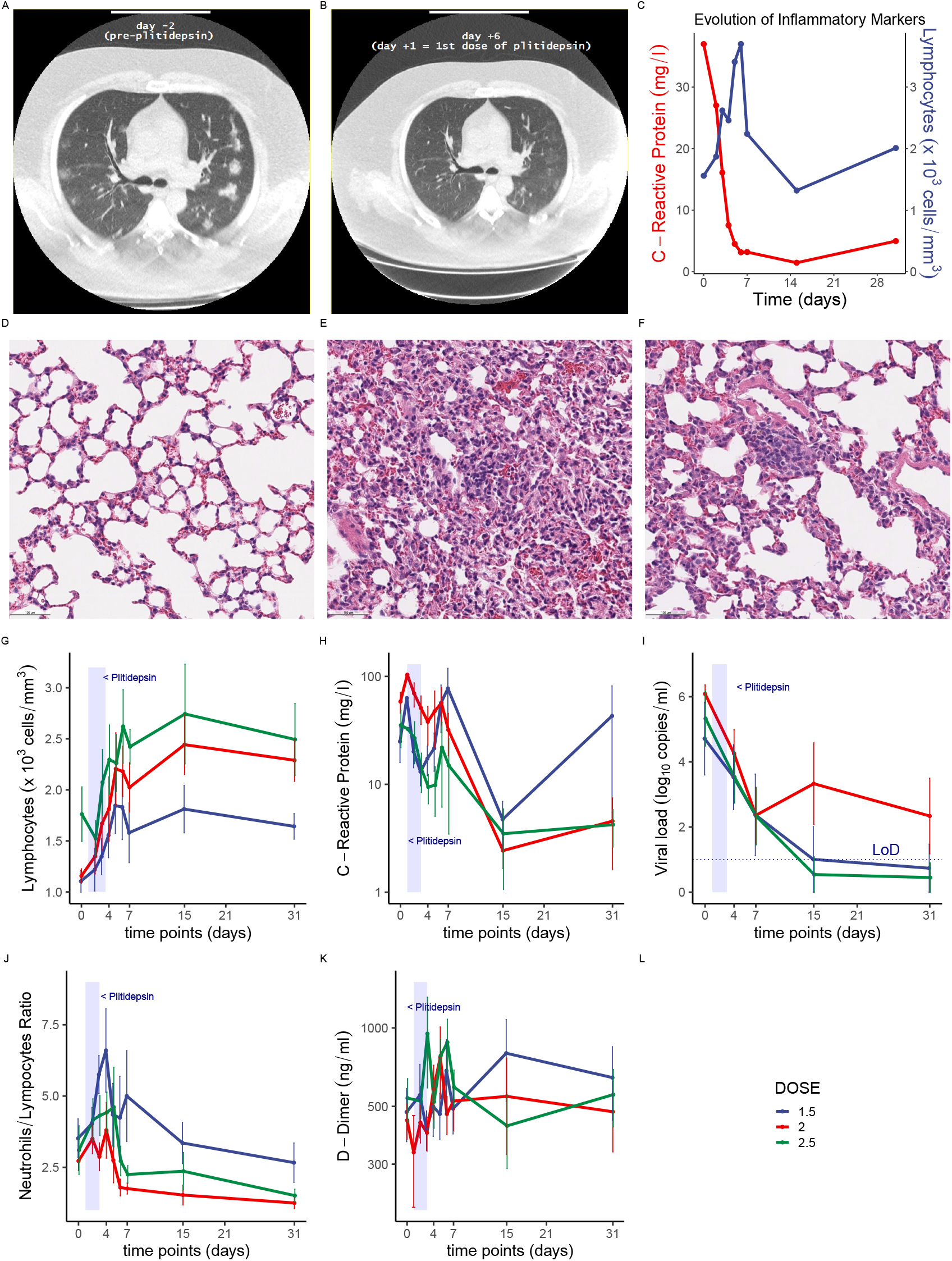
Resolution of inflammatory events post-plitidepsin. (A to C) Evolution of a quadragenarian male patient with moderate COVID-19 who required oxygen supplementation at baseline, treated with plitidepsin 2.5 mg/d: (A & B) Baseline CT scan shows multiple bilateral lung infiltrates 2 days before treatment (A) that improve on day 6 after start of plitidepsin (B); (C) Rapid recovery of abnormal C-reactive protein (CRP) and increase in absolute lymphocyte count in the same patient. Patient was discharged at day 7 (day 1 is the date of the 1^st^ dose of plitidepsin). (D to F) Murine model showing lung alveoli from a non-infected mouse (D), alveolar inflammation in a SARS-CoV-2-infected mouse receiving vehicle (E), and alveolar inflammation in a SARS-CoV-2 infected mouse receiving plitidepsin (F). Plitidepsin reduces lung damage, as reported in *(10)* (G to K) Patients receiving plitidepsin showed improvement over time in several parameters related to infection and inflammation, including lymphocyte counts (G), CRP (H), viral load (I), neutrophil to lymphocyte ratio (J), and D-dimers. LoD - Limit of detection

## Discussion

Since late 2019, the world has been coping with a global health threat caused by the novel coronavirus, SARS-CoV-2. More than one year after being officially declared a pandemic, substantial disease burden remains, as the clinical course from severe lung involvement, evolving to respiratory failure continues to be the main cause of death for patients with COVID-19 patients.

Despite worldwide efforts to identify new treatments, as of this publication, no highly effective antiviral therapy against SARS-CoV-2 is yet available. Several strategies have been attempted with limited effect *(26)*. One large collaborative effort systematically mapped the interactome between SARS-CoV-2 proteins and human proteins, identifying several dozens of potentially druggable interactions *(27)*. Notably, the authors highlighted the potent antiviral effects following the inhibition of eEF1A, which had been previously described as the target of plitidepsin *(9)*.

In the set of preclinical studies of this and previous reports, plitidepsin showed strong antiviral activity and a positive therapeutic index in *in vitro* models of SARS-CoV-2 infection, with better performance than other drugs, including remdesivir *(10, 28)*. In our study, regardless of the coronavirus species (HCoV 227E, SARS-CoV, SARS-CoV-2), the host cells, or the quantifying method used, highly consistent results were obtained, with the IC_50_ of plitidepsin always being in the nanomolar range. Notably, a similar *in vitro* antiviral effect was induced by plitidepsin against the B.1.1.7 variant of SARS-CoV-2 (firstly detected in the United Kingdom), which is known to bear several mutations affecting the viral spike protein that facilitates viral entry through its interaction with the human ACE2 receptor *(29)*.

Using a drug-resistant mutant host factor, White *et al*. demonstrated that the potent *in vitro* activity of plitidepsin against SARS-CoV-2 was mediated through eEF1A inhibition *(10)*. In addition, they also showed strong antiviral activity *in vivo*, characterized by a significant reduction in the viral load in lungs, as well as a clear reduction of alveolar and peribronchial inflammation. In addition, their data also supported a 3-day schedule, as the one used in this clinical study*(10)*.

Treatment with plitidepsin was well tolerated, with most adverse events being mild and transient in nature. There was no relevant hemotoxicity observed in this study. The proportion of laboratory abnormalities was consistent with the expected in COVID-19 patients. *(30)*

The death rate in our study was 6.7%, and was related to COVID-19. A recently published meta-analysis on 33 studies on COVID-19 (totaling 13,398 patients, excluding critical care-only studies), estimated that in hospitalized patients the mortality rate was 11.5% (95% CI: 7.7 - 16.9%)*(31)*. Published data from large retrospective studies on patients admitted into Spanish hospitals report death rates between 20-28% *(32-36)*. It should be noted, however, that these analyses were performed on data extracted from the first epidemic wave, and APLICOV-PC was run during the second wave, which might account in differences in the availability of health resources, learning curve, and baseline severity of hospitalized patients.

Efficacy data gathered from this clinical trial were in agreement with the preclinical antiviral activity of plitidepsin described above. Patients treated with plitidepsin showed rapid reduction in viral load (compared to their baseline value), reduction in biomarkers associated to inflammatory processes, and improvements in pneumonia. Each of these outcomes very likely contributed to mitigating disease progression and leading to an earlier discharge from the hospital.

The antiviral mechanism of plitidepsin may represent significant advantages in the treatment of COVID-19. Specifically, the likelihood of developing treatment-resistant SARS-CoV-2 strains seems to be remote given that plitidepsin does not directly target a viral component. Therefore, SARS-CoV-2 variants that carry mutations to viral components may be equally sensitive to plitidepsin treatment. Morevoer, patients treated with plitidepsin showed a rapid reduction in viral load by RT-PCR. This result needs to be confirmed in larger clinical trials that include a placebo control.

Elevation of inflammation markers such as C-reactive protein is associated with an increased risk of disease severity and mortality *(37, 38)*. In this regard, it is noteworthy that in patients with moderate COVID-19 at baseline, intra-patient variations of inflammation markers trended favorably with higher doses of plitidepsin, which may suggest a drug-effect. These observed changes in inflammatory biomarkers may partly explain the rapid clearance of lung infiltrates observed in chest imaging (Figure 4AB), and is in line with the preclinical observations reported by White *et al*. that treatment with plitidepsin prevents severe lung inflammation. *(10)* (Figure 4CDE)

Translational research on chronic lymphocytic leukemia has identified that plitidepsin can induce cytotoxicity in monocytes at nanomolar concentrations that have little effect in normal lymphocytes *(39)*. Monocytes and macrophages may be infected by SARS-CoV-2, which results in an impairment of the adaptive immune responses against the virus, virus spread, and local tissue inflammation, mediated by the production of large amounts of pro-inflammatory cytokines and chemokines *(40)*. We hypothesize that plitidepsin, besides acting as an antiviral agent, may also modulate immune response by its effects on monocytes/macrophages.

Plitidepsin was hypothesized to hold potential benefits against corona viruses, and therefore SARS-COV-2. This hypothesis was first successfully tested in various *in vitro* work. This led to *in vivo* tests which again demonstrated high potency in inhibiting SARS-CoV-2. We now report a proof-of-concept clinical study, showing the safety of administering plitidepsin at the doses and duration described, and supporting a therapeutic benefit of the treated patients. Nevertheless, our study has several limitations, including the small number of patients evaluated, the large variability, and the lack of a control group. These characteristics are likely limiting the observation of evident dose-response effects. An international controlled phase 3 trial exploring the efficacy and safety of plitidepsin in hospitalized patients with moderate COVID-19 (NEPTUNO; NCT04784559) has already received initial regulatory authorization in UK and several other European countries, the latter through a Voluntary Harmonisation Procedure [VHP1842 (VHP2021019)]. In summary, we have integrated preclinical and clinical studies on the use of plitidepsin to treat SARS-CoV-2 and other coronavirus infections, and generated promising patient data supporting the launching of phase 3 clinical studies to demonstrate the efficacy of treatment with plitidepsin in moderate COVID-19 patients.

## Data Availability

Plitidepsin is available from PharmaMar for noncommercial use under an MTA. All relevant data are included within this manuscript and all materials other than plitidepsin are readily available upon request from the corresponding authors.

## Acknowledgments

We are indebted to the women and men that gave their consent to participate in this study, and to their relatives, for understanding their decision in these exceptional circumstances. We would like to thank Pascal Besman for his input, Timothy Silverstein for providing editorial support, and Lorena Martin Peña for technical and secretarial assistance. The full clinical research teams at each one of the participating sites have played an instrumental role. We recognize hard work and commitment done by Paz Cañadas, Daniel Sánchez-Brualla and Carlota Costa from Synlab Diagnósticos Globales, S.A.U. as central laboratory in charge of viral load characterization. We appreciate the collaboration with Boryung Pharmaceuticals in the first *in vitro* study on the activity of plitidepsin in SARS-CoV-2 infection models. We thank R. Albrecht for support with the BSL3 facility and procedures at the Icahn School of Medicine at Mount Sinai, New York.

## Author contributions

J.F.V., P.L., V.E., R.P., P.G.V., L.F.O., M.T., J.F., R.V., J.B., B.C., J.A., D.C., N.C., L.P., P.G., A.M., D.A. had the role of clinical investigators and participated in patient selection, informed consent, protocol treatment and procedures, local team coordination.

J.B., V.E., J.F. had the role of Coordinating Principal Investigators for APLICOV-PC Clinical Study.

L.E., I.S., S.Z., J.R., N.I.U., S.Y., R.R, K.M.W., N.J.K., L.Z.A., M.B., K.O., A.K.R., L.G.T., C.J., G.J.T., A.G.S conducted cardinal non-clinical research that have either been a guide in the design and execution of this Clinical Trial or are strongly linked to further clinical development.

S.F., M.J.P., B.R., B.S., A.N., J.G., P.G.V, P.A., R.L. were involved in study design, therapeutic intervention modeling, protocol edit and amendments, follow-up and data analysis and coordinators of external translations and studies.

J.M.J., J.A.L.M., J.M.F.S. intervened in the hypothesis, design of the therapeutic model, oversight and design of the statistical analysis and data visualization.

Final study report coordinated by J.A.L.M. and J.M.J.

First draft of the article was produced by J.A.L-M and J.M.J. Major reviewers were P.A, S.F., J.M.F-S, V.E., J.F.V, N.J.K. and A.G-S

The clinical study was conducted within the frame of good clinical practice; Ápices Soluciones, S.L. acted as the Contract Research Organization, in charge of operations, logistics, data monitoring, and data management.

## Supplementary Materials

Materials and Methods

Figures S1-S6

Tables S1-S7

References (1-11)

