## Supplementary information for "Plitidepsin has a positive therapeutic index in adult patients with COVID-19 requiring hospitalization"

**This PDF file includes:**

Plitidepsin – COVID-19 Study Group
Materials and Methods
Figures S1-S6
Tables S1-S7
References (1-11)

**Plitidepsin – COVID-19 Study Group (Alphabetical List)**


| **Co-Author Name** | **Affiliation** |
|---|---|
| Adolfo García-Sastre | Department of Microbiology, Icahn School of Medicine at Mount Sinai, New York, NY, USA.<br>Global Health Emerging Pathogens Institute, Icahn School of Medicine at Mount Sinai, New York, NY, USA.<br>Department of Medicine, Division of Infectious Diseases, Icahn School of Medicine at Mount Sinai, New York, NY, USA.<br>Tish Cancer Institute, Icahn School of Medicine at Mount Sinai, New York, NY, USA. |
| Alfonso Monereo | Internal Medicine Department, Hospital Universitario de Getafe, Madrid, Spain |
| Alvaro Belgrano | PharmaMar - Statistics Unit. Colmenar Viejo, Madrid, Spain. |
| Ana Tercero | PharmaMar - Medical Affairs Unit. Colmenar Viejo. Madrid, Spain. |
| Angelines Barroso | PharmaMar - Medical Affairs Unit. Colmenar Viejo. Madrid, Spain. |
| Ann-Kathrin Reuschl | Division of Infection and Immunity, University College Londo, London, WC1E 6BT, United Kigdom. |
| Antonio Nieto | PharmaMar - Statistics Unit. Colmenar Viejo, Madrid, Spain. |
| Bárbara García | PharmaMar - Medical Affairs Unit. Colmenar Viejo, Madrid, Spain. |
| Beatriz de Rivas | PharmaMar - Medical Affairs Unit. Colmenar Viejo, Madrid, Spain. |
| Belén Sopesén | PharmaMar Virology Unit. Colmenar Viejo, Madrid, Spain.<br>Sylentis, S.A.U., Tres Cantos, Madrid, Spain.<br>Biocross, S.L., Valladolid, Spain. |
| Bonaventura Clotet | Head of Infectious Diseases Department, Director of the Research Lab, IrsiCaixa, Barcelone, Spain. Professor of the UAB and the UVIC-UCC, Barcelone, Spain. |
| Clare Jolly | Division of Infection and Immunity, University College Londo, London, WC1E 6BT, United Kigdom. |
| Cora Loste | Infectious Diseases Department, Hospital Universitari Germans Trias i Pujol (Barcelona, Spain). |
| Daniel Carnevali | Hospital Universitario Quironsalud, Madrid, Spain<br>Universidad Europea, Madrid, Spain. |
| Daniel Abad | Internal Medicine Department, Hospital Universitario de Getafe, Madrid, Spain.<br>European University of Madrid. Spain. |
| Daniel Perez-Zsolt | IrsiCaixa AIDS Research Institute (Barcelona, Spain). |
| Ester Ballana | IrsiCaixa AIDS Research Institute (Barcelona, Spain). |

| Co-Author Name | Affiliation |
| --- | --- |
| Gemma Lladós | Infectious Diseases Department, Hospital Universitari Germans Trias i Pujol (Barcelona, Spain). |
| Greg J. Towers | Division of Infection and Immunity, University College Londo, London, WC1E 6BT, United Kigdom |
| Hervé Dhellot | PharmaMar - Pharmacovigilance Unit. Colmenar Viejo. Madrid, Spain. |
| Isabel Sola | Department of Molecular and Cell Biology, Centro Nacional de Biotecnología (CNBCSIC), Madrid, Spain. |
| Ivette Casafont | Infectious Diseases Department, Hospital Universitari Germans Trias i Pujol (Barcelona, Spain). |
| Javier Gómez | PharmaMar - Statistics Unit. Colmenar Viejo, Madrid, Spain. |
| Jesús Fortún | Hospital Universitario Ramón y Cajal, Madrid, Spain. |
| Joaquim Segalés | IRTA-CReSA (Barcelona, Spain) |
| Jon Cendoya | PharmaMar Virology Unit. Colmenar Viejo. Madrid, Spain. |
| Jordana Muñoz-Basagoiti | IrsiCaixa AIDS Research Institute (Barcelona, Spain). |
| Jordi Rodon | IRTA, Centre de Recerca en Sanitat Animal (CReSA, IRTA-UAB), Campus de la UAB, Bellaterra, Spain |
| Jose A. López-Martín | PharmaMar Virology Unit. Colmenar Viejo. Madrid, Spain. |
| José Barberán | Hospital Universitario HM Montepríncipe, Madrid, Spain. Facultad de Medicina San Pablo CEU, Madrid, Spain. |
| José F. Varona | Departamento de Medicina Interna, Hospital Universitario HM Montepríncipe, HM Hospitales, Madrid, Spain.<br>Facultad de Medicina, Universidad San Pablo-CEU, Madrid, Spain. |
| Jose M. Fernández-Sousa | PharmaMar, S.A., Colmenar Viejo, Madrid, Spain. |
| José M. Jimeno | PharmaMar Virology Unit. Colmenar Viejo. Madrid, Spain. |
| José Ramón Santos | Infectious Diseases Department, Hospital Universitari Germans Trias i Pujol (Barcelona, Spain). |
| Júlia Vergara-Alert | IRTA-CReSA (Barcelona, Spain) |
| Julio Ancochea | Hospital Universitario La Princesa, Madrid, Spain.<br>Centro de Investigación en Red de Enfermedades Respiratorias (CIBERES), Instituto de Salud Carlos III (ISCIII), Madrid, Spain.<br>Universidad Autónoma de Madrid. |
| Kirsten Obernier | Quantitative Biosciences Institute (QBI), San Francisco, CA 94158, USA.<br>J. David Gladstone Institutes, San Francisco, CA 94158, USA. |

| **Co-Author Name** | **Affiliation** |
| --- | --- |
| | QBI, Coronavirus Research Group (QCRG), San Francisco, CA 94158, USA.<br>Department of Cellular and Molecular Pharmacology, University of California, San Francisco, CA 94518, USA. |
| Kris M. White | Department of Microbiology, Icahn School of Medicine at Mount Sinai, New York, NY, USA.<br>Global Health Emerging Pathogens Institute, Icahn School of Medicine at Mount Sinai, New York, NY, USA |
| Laura Soldevila | Infectious Diseases Department, Hospital Universitari Germans Trias i Pujol (Barcelona, Spain). |
| Lola Castro | PharmaMar - Medical Affairs Unit. Colmenar Viejo. Madrid, Spain. |
| Lorena Zuliani-Alvarez | Quantitative Biosciences Institute (QBI), San Francisco, CA 94158, USA.<br>J. David Gladstone Institutes, San Francisco, CA 94158, USA.<br>QBI, Coronavirus Research Group (QCRG), San Francisco, CA 94158, USA.<br>Department of Cellular and Molecular Pharmacology, University of California, San Francisco, CA 94518, USA. |
| Lourdes Mateu | Infectious Diseases Department, Hospital Universitari Germans Trias i Pujol (Barcelona, Spain). |
| Lourdes Porras | Internal Medicine, Hospital General de Ciudad Real, Ciudad Real, Spain. |
| Lucía Bailón | Infectious Diseases Department, Hospital Universitari Germans Trias i Pujol (Barcelona, Spain). |
| Lucía Fernández de Orueta | Internal Medicine Department, Hospital Universitario de Getafe, Madrid, Spain.<br>European University of Madrid. Spain. |
| Lucía Gutiérrez-Chamorro | IrsiCaixa AIDS Research Institute (Barcelona, Spain). |
| Lucy G. Thorne | Division of Infection and Immunity, University College Londo, London, WC1E 6BT, United Kigdom. |
| Luis Enjuanes | Department of Molecular and Cell Biology, Centro Nacional de Biotecnología (CNB-CSIC), Madrid, Spain. |
| María José Pontes | PharmaMar - Medical Affairs Unit. Colmenar Viejo. Madrid, Spain. |
| María Luisa Ramírez | PharmaMar Virology Unit. Colmenar Viejo. Madrid, Spain. |
| Mehdi Bouhaddou | Quantitative Biosciences Institute (QBI), San Francisco, CA 94158, USA.<br>J. David Gladstone Institutes, San Francisco, CA 94158, USA. |

| Co-Author Name | Affiliation |
| --- | --- |
| | QBI, Coronavirus Research Group (QCRG), San Francisco, CA 94158, USA.<br>Department of Cellular and Molecular Pharmacology, University of California, San Francisco, CA 94518, USA. |
| Miguel Torralba | Internal Medicine, Guadalajara University Hospital, Spain<br>University of Alcala, Madrid, Spain. |
| Miriam Ramírez | PharmaMar Virology Unit. Colmenar Viejo. Madrid, Spain. |
| Nevan J. Krogan | Department of Microbiology, Icahn School of Medicine at Mount Sinai, New York, NY, USA.<br>Quantitative Biosciences Institute (QBI), San Francisco, CA 94158, USA.<br>J. David Gladstone Institutes, San Francisco, CA 94158, USA.<br>QBI, Coronavirus Research Group (QCRG), San Francisco, CA 94158, USA.<br>Department of Cellular and Molecular Pharmacology, University of California, San Francisco, CA 94518, USA. |
| Noemí Cabello | Infectious Diseases Department, San Carlos University Hospital. Madrid Spain. |
| Nuria Izquierdo-Useros | IrsiCaixa AIDS Research Institute, 08916, Badalona, Spain.<br>Germans Trias i Pujol Research Institute (IGTP), Can Ruti Campus, 08916, Badalona, Spain. |
| Pablo Avilés | PharmaMar - Preclinical Unit. Colmenar Viejo, Madrid, Spain |
| Pablo Guisado-Vasco | Internal Medicine Department.<br>Hospital Universitario Quironsalud, Madrid, Spain<br>Universidad Europea, Madrid, Spain. |
| Paloma Gijón | Clinical Microbiology and Infectious Diseases Department,<br>Hospital General Universitario Gregorio Marañón<br>Instituto de Investigación Sanitaria Gregorio Marañón, Madrid, Spain. |
| Patricia Girón de Velasco | PharmaMar Virology Unit. Colmenar Viejo. Madrid, Spain. |
| Pedro Landete | Hospital Universitario de La Princesa. Madrid, Spain.<br>Universidad Autónoma de Madrid, Madrid, Spain. |
| Rafael Fernández Alonso | PharmaMar - Medical Affairs Unit. Colmenar Viejo. Madrid, Spain. |
| Roberto Vates | Internal Medicine Department, Hospital Universitario de Getafe, Madrid, Spain. |

| Co-Author Name | Affiliation |
| --- | --- |
| Roger Paredes | Infectious Diseases Department & irsiCaixa AIDS Research Institute, Hospital Germans Trias I Pujol, Badalona, Catalonia, Spain. |
| Romer Rosales | Department of Microbiology, Icahn School of Medicine at Mount Sinai, New York, NY, USA.<br>Global Health Emerging Pathogens Institute, Icahn School of Medicine at Mount Sinai, New York, NY, USA |
| Rubin Lubomirov | PharmaMar - Clinical Pharmacology Unit. Colmenar Viejo, Madrid, Spain |
| Salvador Fudio | PharmaMar - Clinical Pharmacology Unit. Colmenar Viejo, Madrid, Spain |
| Soner Yildiz | Department of Microbiology, Icahn School of Medicine at Mount Sinai, New York, NY, USA.<br>Global Health Emerging Pathogens Institute, Icahn School of Medicine at Mount Sinai, New York, NY, USA. |
| Sonia Extremera | PharmaMar - Statistics Unit. Colmenar Viejo, Madrid, Spain. |
| Sonia Zúñiga | Department of Molecular and Cell Biology, Centro Nacional de Biotecnología (CNBCSIC), Madrid, Spain. |
| Vicente Estrada | Hospital Clínico San Carlos, Madrid, Spain.<br>Universidad Complutense de Madrid, Madrid, Spain. |

### Materials and Methods

Non Clinical Studies

#### a. Enjuanes et al team

***Cells and viral infection:***

Monkey Vero E6 cells were kindly provided by E. Snijder (Leiden University Medical Center, The Netherlands). Human liver-derived Huh-7 cells were kindly provided by R. Bartenschlager (University of Heidelberg, Germany). Cells were cultured in Dulbecco's Modified Eagle Medium (DMEM, Lonza) supplemented with 25 mM HEPES, 10% fetal bovine serum (FBS, HyClone), 2% glutamine, 1% non-essential amino acids (Sigma) and maintained at 37°C in a humidified atmosphere of 5% CO2.

HCoV-229E was kindly provided by V. Thiel (Institute of Virology and Immunology, Switzerland). SARS-CoV virus was rescued from the corresponding infectious cDNAs.*(1)* All experiments with SARS-CoV infectious virus were performed in BSL-3 facilities at CNB-CSIC according to institutional guidelines.

***Experimental design:***

Human Huh-7 cells were infected with HCoV-229E-GFP virus at a MOI (multiplicity of infection) of 0.01. After 8 hpi (hours post infection), medium was replaced by fresh medium containing different plitidepsin concentrations. The presence of fluorescent foci, indicating HCoV-229E-GFP infection, was analyzed at 48 hpi.

SARS-CoV virus stock $2x10^7$ pfu/ml was used. Confluent Vero E6 cells were infected with SARS-CoV at MOI 0.01.After 1 hpi, virus inoculum was retired and medium was replaced by fresh DMEM-HEPES medium with 2% FBS and different concentrations of the compounds:

- Mock infected cells and SARS-CoV infected cells in the absence of DMSO were used as a control to rule out DMSO toxicity effect
- DMSO at the same % present in the compound dilutions was used as a negative control of compound presence
- Plitidepsin and Didemnin B, the most promising compounds in the previous HCoV-229E-GFP screening, were tested at 0.5, 5, 50 nM, 0.1 and 0.5 µM
- PM021473 was used as a control with no effect on CoV infection. It was tested at 0.5, 5, 50 nM, 0.1 and 0.5 µM
- Remdesivir was used as a positive control, tested at 0.5, 5 nM, 0.1, 0.5 and 1 µM

Two independent biological replicates were performed for each condition. At 48 hpi, culture supernatant was collected for virus titration, following standard procedures. (2)

Cells were also collected and total intracellular RNA was purified using RNeasy Mini Kit (Qiagen). Total cDNA was synthesized using 100 ng of total RNA as a template, random hexamers, and a High-capacity cDNA transcription kit (Life Technologies). SARS-CoV gRNA was evaluated using a custom TaqMan assay targeting nsp2 sequence. The human hydroxymethylbilane synthase (HMBS) gene (TaqMan code Hs00609297_m1) was used as a reference housekeeping gene. Data were acquired with a 7500 real-time PCR system (Applied Biosystems) and analyzed with 7500 software v2.0.6. Relative quantifications were performed using the $2^{-\Delta\Delta Ct}$ method. *(3)*

#### b. Rodon et al team

***Cells and viral isolation***:

Vero E6 cells (ATCC CRL-1586) were cultured in Dulbecco's modified Eagle medium, (DMEM; Lonza) supplemented with 5% fetal calf serum (FCS; EuroClone), 100 U/mL penicillin, 100 µg/mL streptomycin, and 2 mM glutamine (all ThermoFisher Scientific).

SARS-CoV-2 was isolated from a nasopharyngeal swab and its genomic sequence deposited at GISAID repository (http://gisaid.org) with accession ID EPI_ISL_510689 as previously described.*(4)*

***Antiviral activity.***

Plitidepsin was assayed from 100 to 0.005 µM in duplicates as previously detailed.*(4)*

Drug dilutions were added to Vero E6 cells and, immediately after, 20 tissue culture infectious dose 50% ($TCID_{50}$) per well of SARS-CoV-2 were inoculated to 30,000 cells in 200 µl. This viral concentration achieves a 50 % of cytopathic effect 3 days post-infection.

Untreated non-infected cells and untreated virus-infected cells were used as negative and positive controls of infection, respectively. To detect any drug-associated cytotoxic effect, Vero E6 cells were equally cultured in the presence of increasing drug concentrations, but in the absence of virus.

Viral-induced cytopathic or drug-induced cytotoxic effects were measured 3 days post infection, using the CellTiter-Glo luminescent cell viability assay (Promega). Luminescence was measured in a Fluoroskan Ascent FL luminometer (ThermoFisher Scientific).

Cells not exposed to the virus were used as negative controls of infection and were set as 100% of viability to normalize data and calculate the percentage of cytopathic effect. Response curves of compounds were adjusted to a non-linear fit regression model, calculated with a four-parameter logistic curve with variable slope.

#### c. Krogan/Garcia-Sastre et al team:

***Cells culture and drugs*:**

Calu-3 cells (ATCC HTB-55) and Caco-2 cells were a kind gift Dr Dalan Bailey (Pirbright Institute).

Cells were cultured in Dulbecco's modified Eagle Medium (DMEM) supplemented with 10% heat-inactivated FBS (Labtech), 100U/ml penicillin/streptomycin, with the addition of 1% Sodium Pyruvate (Gibco) and 1% Glutamax for Calu-3 and Caco-2 cells. All cells were passaged at 80% confluence. For infections, adherent cells were trypsinized, washed once in fresh medium and passed through a 70 µm cell strainer before seeding at 0.2x10^6 cells/ml into tissue-culture plates. Calu-3 cells were grown to 60-80% confluence prior to infection as described previously. *(5)*

Plitidepsin (PharmaMar), and remdesivir (SelleckChem) were reconstituted in sterile DMSO.

***Viruses:***

SARS-CoV-2 strain BetaCoV/Australia/VIC01/2020 (NIBSC) and SARS-CoV-2 B.1.1.7 (SARS CoV 2 England/ATACCC 174/2020) strain were propagated by infecting Caco-2 cells at MOI 0.01 TCID50/cell, in DMEM supplemented with 10% FBS at 37°C.

Virus was harvested at 72 hours post infection (hpi) and clarified by centrifugation at 4000 rpm for 15 min at 4 °C to remove any cellular debris. Virus stocks were aliquoted and stored at -80 °C.

Virus titers were determined by quantification of SARS-CoV-2 RNA genomes/ml as previously described.*(5)*

***Infection and drug assays:***

Calu-3 and Caco-2 cells were pre-treated with remdesivir or plitidepsin at the indicated concentrations or DMSO control at an equivalent dilution for 2 h before SARS-CoV-2 infection.

Caco-2 and Calu-3 cells were infected at $1x10^3$ E copies per cell, equivalent to an MOI of 0.01 TCID50 per cell (as titered on Vero.E6). Inhibitors were maintained throughout infection.

Cells were harvested after 24h for analysis and viral infection measured by intracellular detection of SARS-CoV-2 nucleoprotein by flow cytometry. Tetrazolium salt (MTT) assay was performed to verify cell viability. 10 % v/v MTT was added to the cell media and cells were incubated for 24 h at 37ºC. Cells were lysed with 10% SDS, 0.01M HCl and the formation of purple formazan was measured at 620nm.

***Flow cytometry:***

For flow cytometry analysis, adherent cells were recovered by trypsinization and washed in PBS with 2mM EDTA (PBS/EDTA). Cells were stained with fixable Zombie NIR Live/Dead dye (Biolegend) for 6 min at room temperature. Excess stain was quenched with FBS-complemented DMEM. Cells were fixed in 4% PFA prior to intracellular staining.

For intracellular detection of SARS-CoV-2 nucleoprotein, cells were permeabilized for 15 min with Intracellular Staining Perm Wash Buffer (BioLegend). Cells were then incubated with 1µg/ml CR3009 SARS-CoV-2 cross-reactive antibody (a kind gift from Dr. Laura McCoy) in permeabilization buffer for 30 min at room temperature, washed once and incubated with secondary Alexa Fluor 488-Donkey-anti-Human IgG (Jackson Labs). All samples were acquired and analysed on a NovoCyte 3005 Flow Cytometer System (Agilent).

#### d. <u>Boryung Pharmaceutical.*(6)*</u>

***Virus and cells***

Both Vero (ATCC CCL-81) and Calu-3 cells (ATCC HTB-55) were purchased from the American Type Culture Collection (ATCC). SARS-CoV-2 (βCoV/KOR/KCDC03/2020) was provided by Korea Centers for Disease Control and Prevention (KCDC) and was propagated in Vero cells. Viral titers were determined by plaque assays in Vero cells.

***Reagents.***

Plitidepsin (batch#: 16 D19) and ampoule (batch#: 60108) were provided by Boryung Pharmaceutical. Anti-SARS-CoV-2 N protein antibody was purchased from Sino Biological lnc. (Beijing, China). Alexa Fluor 488 goat anti-rabbit [IgG (H+L) secondary antibody and Hoechst 33342 were purchased from Molecular Probes.

***Drug treatment, infection and immunofluorescence staining***

Two milligrams of plitidepsin was dissolved in either 1,801.3 µl of DMSO or ampoule at a final concentration of 1 mM and a two-fold dilution series was made with 20-points. Vero cells were seeded at l .2x 10 4 cells per well in DMEM supplemented with 2% FBS and I X Antibiotic- Antimycotic solution (Gibco) in black, 384-well, µClear plates (Greiner Bio-One), 24 hours before plitidepsin treatment and virus infection. Calu-3 cells were seeded at 2.0 x 104 cells per well in EMEM supplemented with 10 % FBS and 1X Antibiotic- Antimycotic solution (Gibco) in black, 384-well, µClear plates (Greine r Bio-One), 24 hours before plitidepsin treatment and virus infection. Twenty-point plitidepsin dilution series generated above was added to Vero or Calu-3 cells with the highest concentration at 5 µM. After 1 hour, the plates were transferred into the BSL-3 containment facility for virus infection. SARS-CoV-2 was added at a multiplicity of infection (MO1) of 0.0125 and 0.1 to the plates for Vero cells and Calu-3 cells, respectively.

The cells were fixed at 24 hpi with 4 % paraformaldehyde. Anti-SARS-CoV-2 nucleocapsid (N) antibody and Alexa Fluor 488-conjugated goat anti-rabbit IgG antibody were used for immunostaining of viral N protein and Hoechst 33342 were used to stain nuclei of the host cells.

***Image analysis***

The images acquired with Operetta (Perkin Elmer) were analyzed using our in-house Image-Mining (IM) software to quantify cell numbers and infection ratio by counting Hoechst-stained nuclei and viral N protein-expressing cells, respectively. The infection ratio of each well was normalized to the average of infection percentage of infection group (0.5 % DMSO) and mock infection group in each plate. The cell ratio was determined according to the number of cells of each well versus the average number of cells of mock infection in each plate and described as “cell number to mock” in the dose-response curve (DRC) graph. The DRCs of plitidepsin (both. dissolved either in DMSO or in ampoule), was analyzed using the XLfit® equation: Y= Bottom +(Top - Bottom)/(1 + (IC50/X)Hillslope).

The IC50 and 50 % toxicity concentration (CCso) values were calculated from the fitted dose-response curves. Selectivity index (SI) was calculated as CC50/IC50. All IC50 and CC50 values were determined in duplicate experiments. All the experiments were simultaneously.

### Model-based dose justification

***In vitro activity data***

The results from Boryung Pharmaceuticals in infected Vero cells demonstrated a very strong antiviral effect induced by plitidepsin, with half-maximal inhibitory concentration ($IC_{50}$) as low as 3.26 (95%, 2.973-3.585) nM. These were consistent with results from a different cell line, such as Calu-3. Based on the resulting Hill slope (2.082) and according to the following equation,*(7)* a 90% maximal inhibitory concentration ($IC_{90}$) of 9.38 (95%CI, 7.653-11.50) nM was estimated.

$$ICF_{in\ vitro} = \left(\frac{F}{100-F}\right)^{1/H} \times IC50_{in\ vitro},$$

where F is the percentage of response (i.e. 90%), and H is the Hill slope

***Plasma protein binding***

The binding of plitidepsin to plasma proteins was determined *in vitro* at three concentrations (100, 250 and 500 ng/mL) in male rat, dog and human plasma.*(8)*

Rat (CD), Beagle dog and human blood was collected at Aptuit Srl (Verona, Italy) from at least three male donors, and plasma prepared by centrifuging blood at 2000 g, 4°C for 10 minutes. Human plasma was obtained from healthy and fasted male volunteers. Na-heparin was used as the anti-coagulant for all species. All plasma was stored at approximately -20°C and thawed only once, on the day of the experiment.

Plasma protein binding was assessed using Rapid Equilibrium Dialysis (RED) plates pre-loaded with equilibrium dialysis membrane inserts (MWCO ~8 kDa).

Separation of free compound from protein-bound material was achieved by dialysis of the sample through the membrane under appropriate shaking at 37°C.

The suitability of equilibrium dialysis as a method for protein binding determination of plitidepsin was assessed.

Prior to experimentation, the time required to reach equilibrium was determined in male rat and human plasma at one concentration (250 ng/mL) and six selected timepoints (2, 3, 4, 5, 6, 7 h) at 37°C.

Plitidepsin stability assay was assessed in plasma (all species) and PBS (phosphate buffered saline) at two concentrations (100 and 500 ng/mL for plasma; 0.5 and 50 ng/mL for PBS) by comparing plitidepsin concentration in spiked and preincubated plasma prior to and after incubation at 37oC for 5 hours as determined previously.

***Tissue distribution.***

An *in vivo* distribution study was carried out in rats.

PM140064 ($^{14}C_1$-Plitidepsin) was supplied by PharmaMar at a radiochemical purity of 95 % or greater, with no single impurity 3 % or greater. The study was conducted at Aptuit Srl (Verona, Italy). *(9)*

Twenty-four male and twenty-four female Sprague-Dawley rats each received a single IV bolus administration of [$^{14}C$]Plitidepsin at a target dose of 0.2 mg/kg. Following administration, three animals per time-point (0.25, 1, 2, 4, 8, 24, 48 and 72 h) post-dose) were exsanguinated from the abdominal aorta and blood retained. Actual times of bleeding were recorded. In addition to blood, the following tissues were collected: brain, eyeballs, heart, liver, lung, skeletal muscle (quadriceps), fat, kidneys, stomach, skin, small intestine, spleen, thyroid, lymph nodes and testes or ovaries. At the end of the collection period animals will be sacrificed by CO2 asphyxiation and the carcass discarded. Additional animals were bled to obtain control blood/plasma and tissues.

***Estimation of target IC50 and IC90 total plasma concentrations:***

Results from previously described studies show that: a) in human plasma, plasma-protein binding of plitidepsin was estimated at 98%, independent of drug concentration (Table S2), and b) after the administration of a single intravenous bolus dose (0.2 mg/kg) of plitidepsin ($^{14}C$-labelled), a significant increased distribution of radioactivity was found in lung; this preferential distribution led to lung-to-plasma AUC ratio (LPR) of approximately 543-fold, respectively (calculated from Table S3).

The partition coefficient LPR enables the quantification of the total drug concentration in the tissue, and, by assuming similar lung distribution in humans to that observed in rodents and similar fraction of unbound drug in plasma and tissue, unbound exposures in human lung can be further estimated.

Therefore, total plasma concentration (µg/L) of plitidepsin associated with lung exposures above the *in vitro* target concentration $IC_{50}$ and $IC_{90}$, were estimated at 0.33 (95%CI, 0.30-0.37) µg/l and 0.96 (95%CI, 0.78-1.18) µg/l, respectively, according to the equation bellow. This equation considers plasma protein binding and lung-plasma AUC ratio, based on current recommendations.*(10)*

$$\mathrm{ICF}_{\mathrm{total,plasma}} = \frac{\mathrm{ICF}_{total,\,in\,vitro}}{f_{u,\,human} \cdot \, LPR_{rat}}$$

where $ICF_{total,plasma}$ is the total target plasma concentration (µg/l), $ICF_{total,\textit{in vitro}}$ is the concentration (µg/l) used in the *in vitro* experiment, $f_{u,human}$ is the unbound fraction in human plasma, and $LPR_{rat}$ is the lung-plasma AUC ratio in the distribution study in rats.

***Simulation of plitidepsin plasma exposures at the selected dose regimen***

A validated pharmacokinetic population model of plitidepsin*(11)* updated with data from multiple myeloma patients, was used to simulate plitidepsin plasma profiles in typical subjects treated with the selected dose regimen (D1-3 in 1.5 hour infusion), in order to observe whether they would reach the estimated target plasma concentrations with antiviral activity. This model was developed based on plasma and blood concentrations from 549 cancer patients from four phase 1, nine phase 2 and one phase 3 study treated with plitidepsin either as monotherapy or in combination with dexamethasone, at doses ranging from 0.8 to 8.0 mg/m$^2$ as a 1-hour or 24-hour infusion weekly, 3-hour or 24-hour infusion biweekly, or 1-hour infusion daily for 5 consecutive days every three weeks. An open, 3-compartment disposition model with linear elimination and linear distribution from the central compartment to two peripheral compartments was used to describe the PK of plitidepsin in plasma. The model was parameterized in terms of systemic clearance (Cl), central ($V_1$) and peripheral volume of distribution for the shallow ($V_2$) and deep ($V_3$) compartments, and intercompartmental exchange flows for shallow ($Q_2$) and deep ($Q_3$) compartments. The concentration of plitidepsin bound to red blood cells was modeled as a nonlinear function and the plitidepsin blood concentration was estimated according to the following equation*(11)*:

$$C_{blood} = C_{plasma} \cdot (1 - HCT) + \frac{B_{\max} \cdot C_{plasma}}{k_d + C_{plasma}} \cdot HCT$$

where $B_{max}$ corresponds to the maximal plitidepsin concentration bound to blood cells, $k_d$ is the plitidepsin plasma concentration at which the plitidepsin bound to red blood cells is half-maximal, and HCT is the baseline hematocrit of each patient.

Based on the deterministic simulations of plasma concentration-time profiles, flat doses of 1.5, 2.0 and 2.5 mg will be associated to plasma concentrations above IC50 throughout the whole treatment period and will remain above IC90 during most of the administration interval, as depicted in Figure 2, while accumulation after three repeated administrations is minimal.

Clinical proof-of-concept study (APLICOV-PC)

**Title of Study:** Multicenter, Randomized, Parallel and Proof of Concept Study to Evaluate the Safety Profile of Three Doses of Plitidepsin in Patients with COVID-19 Requiring Hospitalization.

**Protocol Number:** APLICOV-PC (APL-D-002-20)

**EudraCT Number:** 2020-001993-31

**clinicaltrials.gov Registry Number:** NCT04382066
https://clinicaltrials.gov/ct2/show/NCT04382066

**Name of Sponsor Company:** PharmaMar, S.A.

**Name of Finished Product:** Plitidepsin

**Name of Active Ingredient** (IUPAC): (S)-N-((R)-1-(((3S,6R,7S,10R,11S,15S,17S,20S,25aS)-10-((S)-sec-butyl)-11-hydroxy-20-isobutyl-15-isopropyl-3-(4-methoxybenzyl)-2,6,17-trimethyl-1,4,8,13,16,18,21-heptaoxodocosahydro-1H-pyrrolo[2,1-f][1,15,4,7,10,20] dioxatetraazacyclotricosin-7-yl)amino)-4-methyl-1-oxopentan-2-yl)-N-methyl-1-(2-oxopropanoyl)pyrrolidine-2-carboxamide

**Coordinating Principal Investigators:** Vicente Estrada, MD, Hospital Clínico Universitario San Carlos (Madrid, Spain); Jesús Fortún Abete, MD, Hospital Ramón y Cajal (Madrid, Spain); José Barberán, Hospital HM Montepríncipe (Madrid, Spain)

**Study period:**
First patient in: 12-May-2020
Last patient follow up: 26-Nov-2020

**Objectives:**

***Primary Objective:***

- Determine the safety and toxicological profile at each dose level, based on
- Frequency of grade ≥ 3 adverse events (AEs) at Days 3, 7, 15 and 31, based on NCI CTCAE v.5.0 criteria;
- Percentage of patients unable to complete treatment and reasons;
- Percentage of patients with AEs and SAEs at Days 3, 7, 15 and 31;
- Change from baseline (Day -1 or Day 1 before administration of plitidepsin) in hematologic and non-hematologic parameters on Days 3, 7 15 and 31;
- Percentage of patients with ECG abnormalities on Days 2, 3, 4, 5, 6, 7, 15 and 31.

***Secondary Objectives:***

- Assess efficacy at each dose level based on
- Change in SARS-CoV-2 viral load from baseline (Day -1 or Day 1 before administration of plitidepsin) measured on Days 4, 7, 15 and 31;
- Time until negative detection of SARS-CoV-2 by PCR;
- Mortality at Days 7, 15 and 31;
- Percentage of patients requiring invasive mechanical ventilation and/or ICU admission at Days 7, 15 and 31;
- Percentage of patients requiring non-invasive mechanical ventilation at Days 7,15 and 31;
- Percentage of patients requiring oxygen therapy at Days 7, 15 and 31.

- Select the recommended dose levels for a phase 2/3 study based on joint evaluation of study data by the Sponsor and the Spanish Agency of Medicine and Medical Devices

***Methodology:*** This was a phase 1, proof-of-concept, multicenter, open-label study in which patients hospitalized for management of COVID-19 infection were enrolled sequentially into three dose groups, including 1.5 mg, 2.0 mg, and 2.5 mg plitidepsin, administered as a 90- minute IV infusion once a day for three consecutive days. The first patients in the 2.0 mg cohort could not be enrolled until the first 3 patients in the 1.5 mg cohort had successfully completed Day 15 assessments (12 days after completing plitidepsin dosing); similarly, the first 3 patients in the 2.5 mg cohort could be not enrolled until the first 3 patients in the 2.0 mg cohort had successfully completed Day 15 assessments (12 days after completing plitidepsin dosing). When multiple dose levels were open for enrollment, patients were randomized 1:1 or 1:1:1 as appropriate.

Continuation of enrollment into a dose level was based on the following criteria:

- If 1 of 3 patients treated at 1.5 mg qdx3 experiences a grade ≥ 3 AE within the 12-day follow-up after administration of the last dose, 3 additional patients will be enrolled in the 1.5 mg cohort and recruitment of the first 3 patients in the 2.0 mg cohort will be initiated. Recruitment into the 2.5 mg cohort will be initiated only after it has been shown that no more than 1 of 3 patients treated at 2.0 mg qdx3 experiences a grade ≥ 3 AE within the 12-day follow-up after administration of the last dose of 2.0 mg.
- If 2 or more patients in a dose cohort experiences a grade ≥ 3 AE, enrollment in the cohort will be placed on hold and the Sponsor will jointly assess with the Spanish Agency of Medicine and Medical Devices whether or not to continue recruitment into that cohort and any higher dose cohorts.
- Intra-patient dose change/increase was not allowed.

Following completion of the first 27 patients, the protocol was amended to add an additional 18 patients, randomized 1:1:1 to each dose cohort, to obtain additional information on treatment tolerance and efficacy.

***Number of Patients***: 45 planned patients

**Diagnosis & Main Criteria for Inclusion:**

***Key inclusion criteria:***

- Signed informed consent prior to initiation of any study-specific procedures
- COVID-19 infection confirmed by real-time reverse transcription polymerase chain reaction (PCR) testing of a nasopharyngeal exudate or sample from the lower respiratory tract
- Patients who require hospitalization for COVID 19
- Onset of COVID-19 infection symptoms no later than 10 day prior to study inclusion
- Men or women (non pregnant) aged ≥ 18 years
- Women of reproductive capacity must have a negative pregnancy test and must be non-lactating
- Women and men with partners of childbearing potential must take effective contraception while on study and for 6 months after the last dose of plitidepsin

***Key Exclusion Criteria:***

- Participation in another clinical trial for COVID-19 infection
- Receiving antivirals, interleukin-6 receptor inhibitors, or immunomodulatory drugs for treatment of COVID-19 infection
- Receiving treatment with chloroquine and derivatives
- Evidence of multi-organ failure
- Requiring support with mechanical ventilation (non-invasive or invasive) at study enrollment

- Laboratory values at screening visit:
    - D-dimer > 4 x upper limit of normal (ULN)
    - hemoglobin (Hb) < 9 g/dL
    - absolute neutrophil count (ANC) < 1000/mm3
    - platelets < 100,000/mm3
    - lymphocytes < 800/mm3
    - aspartate aminotransferase (AST, SGOT) or alanine aminotransferase (ALT, SGPT) > 3 x ULN
    - total bilirubin > 1 x ULN
    - creatinine phosphokinase (CPK) > 2.5 x ULN
    - creatinine clearance < 30 mL/min
    - serum troponin > 1.5 x ULN
- Clinically relevant heart disease (Hew York Heart Association Class >2)
- Clinically relevant arrhythmia or history/presence of QTc interval prolongation ≥ 450 ms
- Neuropathy of any type grade ≥ 2
- Requirement for or treatment with strong inhibitors or inducers of CYP3A4
- Hypersensitivity to the active substance or any of the excipients (macrogolglycerol ricinoleate and ethanol)
- Patients who for any reason should not be included in the study according to the evaluation of the research team

**Test Product, Dose and Mode of Administration, Batch Number(s):** Plitidepsin was administered as an IV infusion over 90 minutes on Days 1, 2 and 3 at dose levels of 1.5, 2.0 and 2.5 mg. A single batch (16D19) was administered to all patients (expiry date 04/21).

**Duration of Treatment:** Treatment for 3 consecutive days (Days 1-3), with follow-up for safety and efficacy through Day 31

**Treatment and co-medications:** See Figure S2.

**Criteria for evaluation:**

***Safety:***

- Adverse events evaluated at baseline, Days 1-7, 15 and 31, and graded in terms of severity (NCI CTCAE v.5.0) and relationship to study drug
- Vital signs (temperature, blood pressure, heart rate, respiratory rate, SpO2 including oxygenation method) every 8 hours (or once each nursing shift) during hospitalization, then at protocol-specified visits through Day 31
- Hematology (CBC) at baseline, Days 1-7, 15 and 31
- Coagulation parameters including D-dimer at baseline, Days 1-7, 15 and 31
- Serum chemistry (ALT, AST, GGT, total bilirubin, alkaline phosphatase, LDH, CPK, ferritin, troponin) and C-reactive protein at baseline, Days 1-7, 15 and 31
- 12-lead ECGs for PR and QT intervals at baseline, Days 1-7, 15 and 31

***Efficacy:***

- Quantitative PCR test by central lab on nasopharyngeal exudate or sample from lower respiratory tract within 48 hours prior to first administration of plitidepsin and on Days 4, 7, 15 and 31 (performed to assess viral load)
- Percentage of patients requiring oxygen therapy, non-invasive mechanical ventilation, invasive mechanical ventilation and/or ICU admission, and dead on Days 7, 15 and 31

**Statistical methods:**

Analysis of safety data was conducted on all patients who received at least one dose of study drug and the results were reported using descriptive statistics.

Analysis for preliminary evidence of efficacy was performed on all patients who had efficacy assessments at baseline and at least one subsequent time on treatment.

Post-hoc analyses were conducted to (1) assess disease severity at baseline (mild, moderate, severe COVID-19 by FDA criteria) and (2) assess covariates associated with efficacy endpoints, including hospital discharge by Day 15 or Day 8. Safety and efficacy outcomes also were assessed by age group (<65 years vs ≥65 years) and efficacy outcomes were assessed by disease severity at baseline. Safety outcomes were also assessed before and after the implementation of amendment AR9.

Continuous and categorical endpoints for the treatment effect are reported. The differences in proportions are reported with 95% confidence intervals (CIs). Formal statistical testing between subgroups were not pre-defined due to the reduced numbers in each subgroup. All statistical analyses and plots were carried out using SAS software 9.4 (SAS Institute, Cary, NC, USA) and R 4.0.3.

**SARS-CoV-2 quantification**

Centralized assessments of nasopharyngeal samples were performed at SYNLAB (Esplugues de Llobregat, Spain).

Viral RNA extraction from the samples was done using the Maxwell HT Viral TNA (AX2340) Promega, on the Hamilton automated extraction platform. Detection of nCoV2019 was carried out with the kit: TaqPath TM COVID-19 RT-PCR KIT (A48102)) Thermofisher.

The positivity / negativity criterion was based on the following table:

| ORF1ab gen ≤37 CT value | N gen ≤37 CT value | S gen ≤37 CT value | Internal Control ≤30 CT value | SARS-CoV-2 RESULT |
|---|---|---|---|---|
| **Undetected** | Undetected | Undetected | Invalid | Repeat plate |
| **Undetected** | Undetected | Undetected | Positive | Undetected |
| **Just a target gen is detected** | | | Positive/Negative | Repeat plate |
| **Two or more targets detected** | | | Positive/Negative | Positive |

Each sample was analyzed in triplicate, and the CT value obtained was extrapolated to the standard curve, so that a value of viral load is obtained per reaction for said sample.

The quantification, in triplicate, of the viral load of the patient sample was carried out at each of the 5 timepoints.

For quantification, in each sample plate, 8 points, in triplicate (24 PCR in total), of the plasmid IDT 2019 nCov Kit CDC EUA of known viral load was used to generate the standard line on which to extrapolate the results of the samples of the trial patients.

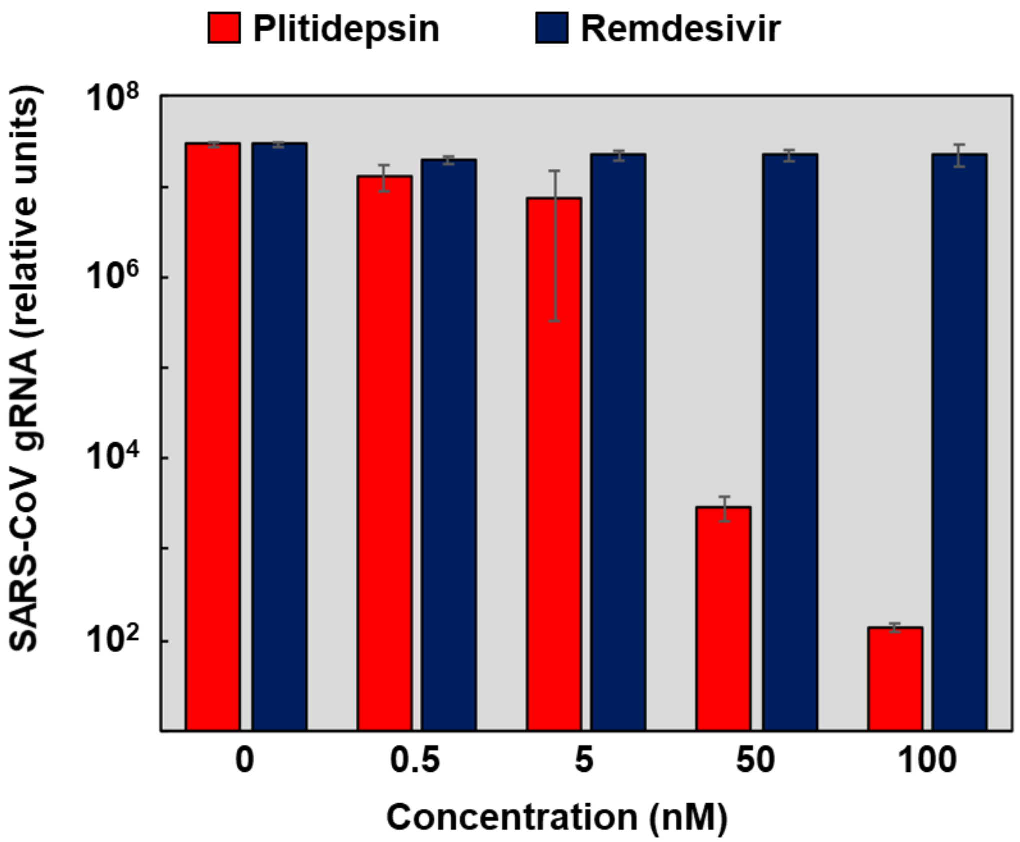


**Figure S1. Comparative effects of plitidepsin in viral gRNA synthesis (Enjuanes et al)**
A significant $10^4$-fold decrease in gRNA accumulation was observed in cells treated with Aplidin 50 nM, respect to DMSO. Remdesivir, used as a positive control, had no effect on virus RNA synthesis.

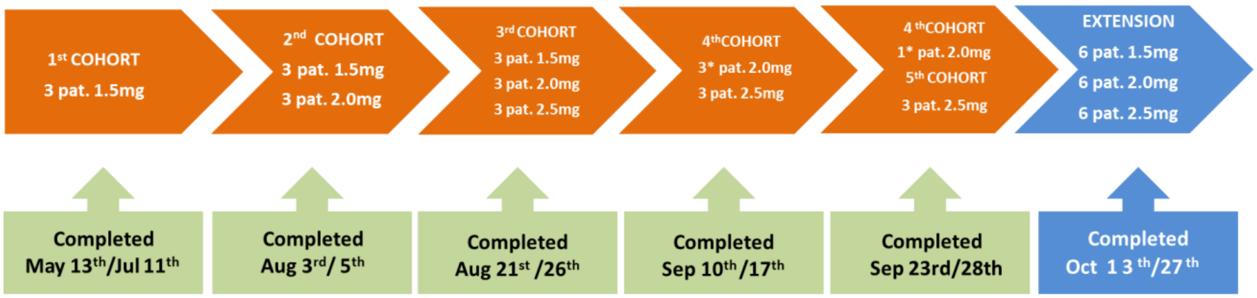


**Figure S2.APLICOV-PC Study flow**

46 patients recruited - 1 patient withdrew consent before initiating study procedures
45 patients treated:

- 15 patients on 1,5 mg/day x 3 days
- 15 patients on 2,0 mg/day x 3 days
- 15 patients on 2,5 mg/day x 3 days

44 patients completed treatment

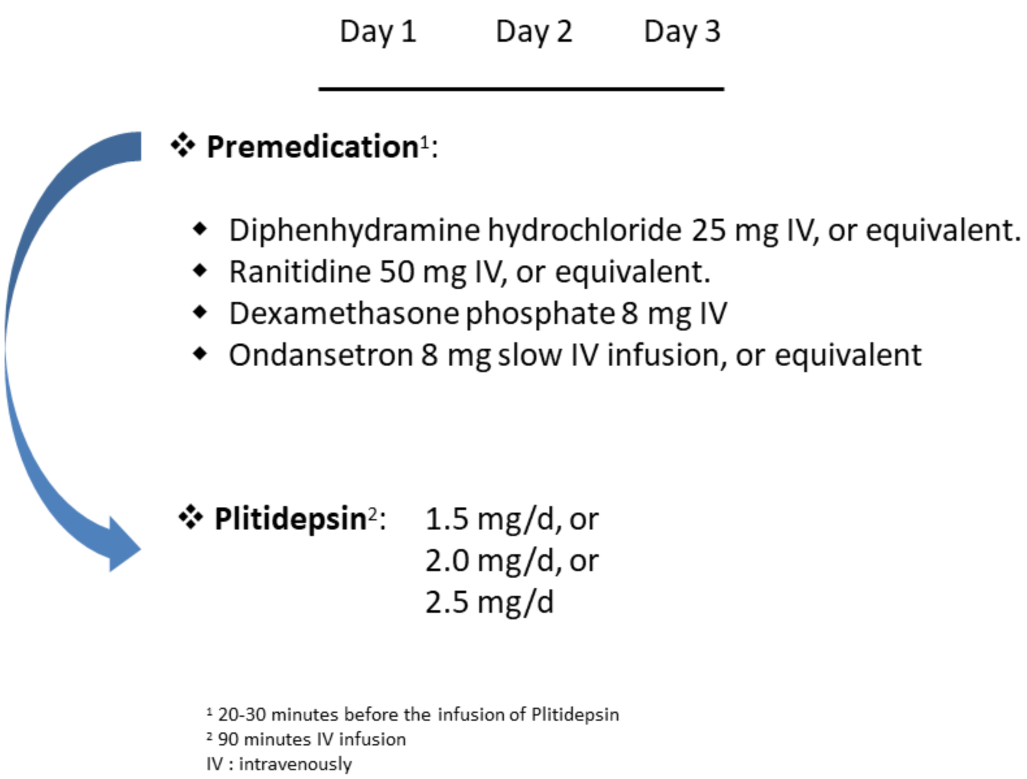


**Figure S3. APLICOV-PC: Protocol Treatment**

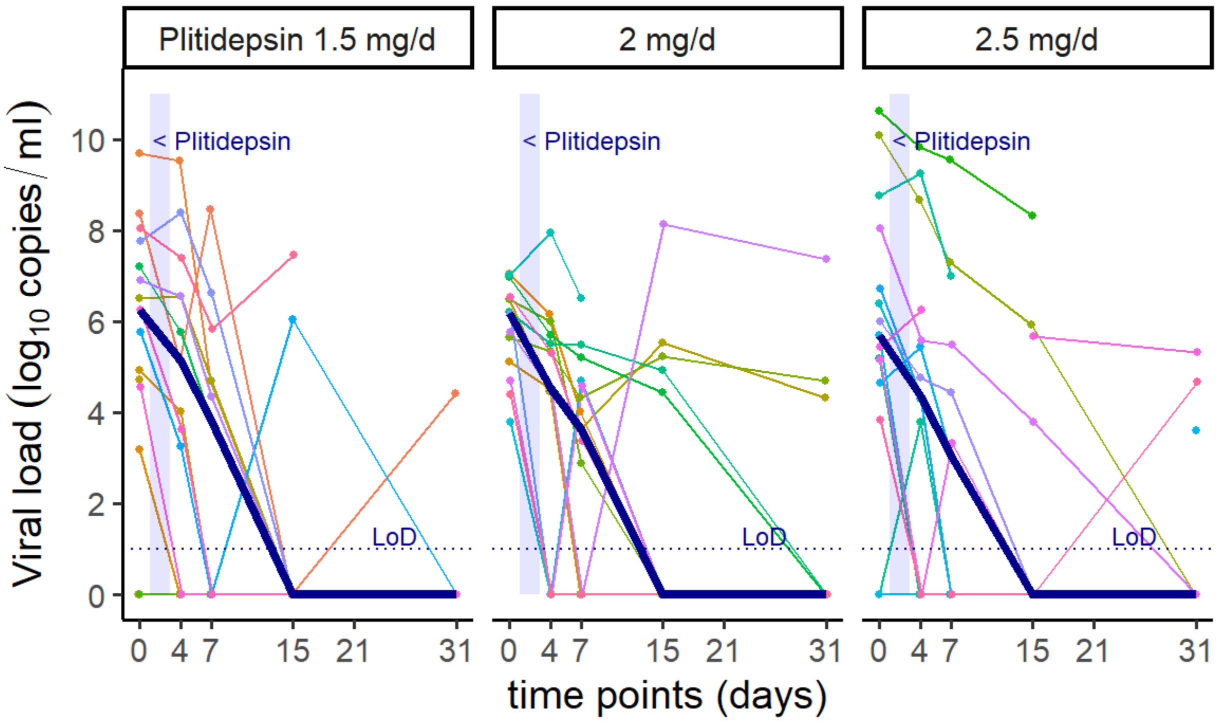


**Figure S4. Viral load kinetics per dose cohort**

Viral load assessed by quantitative RT-PCR from nasopharyngeal samples.
Shadowed area represents treatment with plitidepsin on Days 1 to 3.
Each color line represents one patient.
Bold line represents median values.

**LoD:** Limit of detection

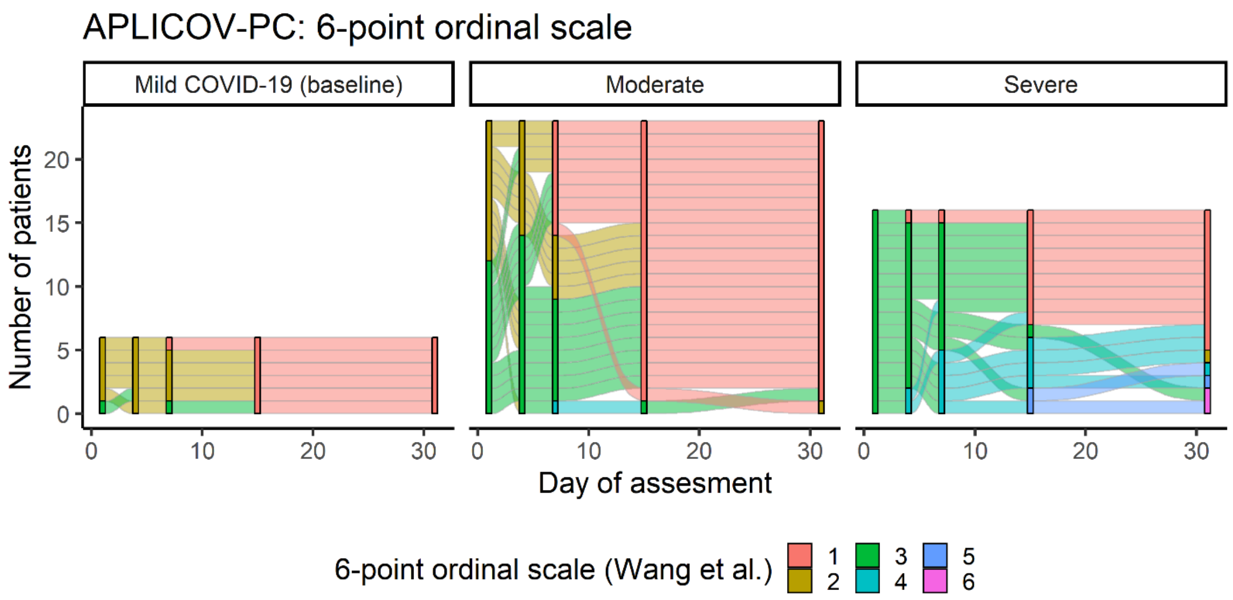


**Figure S5. Individual time-variation of the assessment of a 6-categories ordinal scale (Wang et al.) according to the severity of the disease at baseline.**

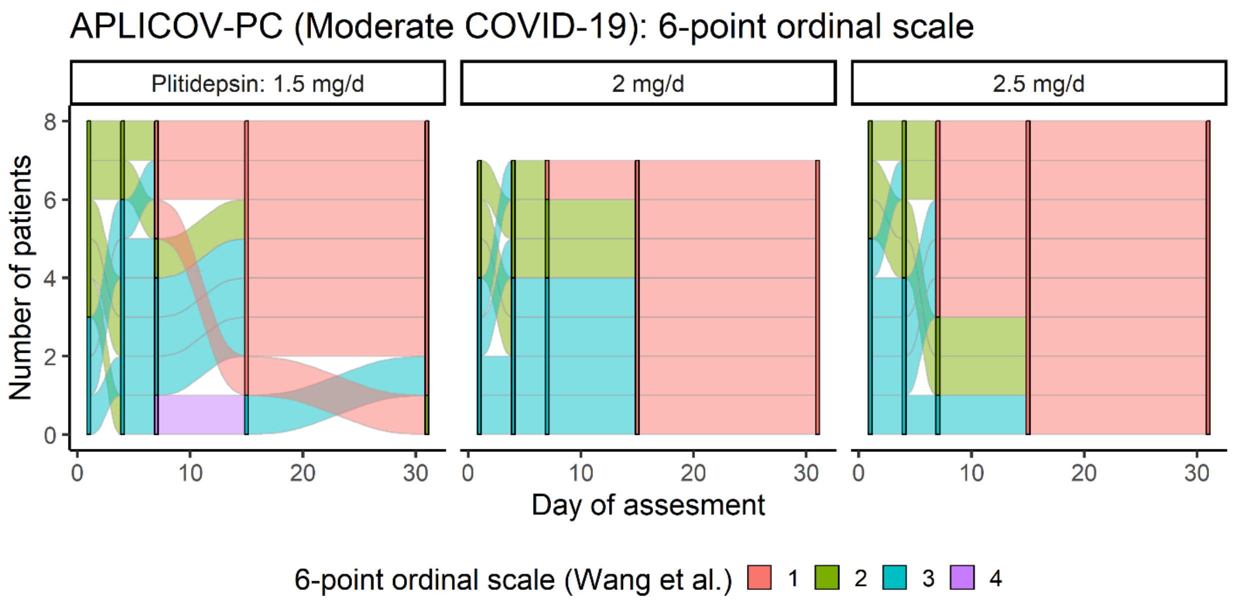


**Figure S6. Individual time-variation of the assessment of a 6-categories ordinal scale (Wang et al.) in patients with moderate COVID-19 at baseline, according to the administered dose of plitidepsin.**

**Table S1. Comparative effects of plitidepsin in SARS-CoV virus titers. (Enjuanes et al)**

| COMPOUND | CONCENTRATION | TITER, pfu/ml |
|---|---|---|
| DMSO | -- | $(1.4 \pm 0.3) \times 10^6$ |
| Plitidepsin | 5 nM | $(2.3 \pm 0.6) \times 10^6$ |
| | 50 nM | $\leq 1 \times 10^3$ |
| Remdesivir | 1 µM | $(4.4 \pm 0.5) \times 10^6$ |

A $10^3$-fold decrease in SARS-CoV titers was observed in the presence of Aplidin 50 nM. Remdesivir, used as a positive control, had no effect on virus titer, although an $EC_{50}$ of 74 nM for SARS-CoV was previously determined in human airway epithelial (HAE) cell cultures (Agostini et al, 2018). Since Remdesivir requires metabolism by cell factors (Cho et al, 2012), it is possible that these factors are not conserved in Vero E6 cells.

**Table S2. Extent of In Vitro Binding of Plitidepsin to Plasma Proteins in Human Determined by Equilibrium Dialysis.*(8)***

| Nominal | Concentration (ng/mL) Actual ($t_0$) | Dialyzed Plasma | Dialyzed PBS | % Unbound | % Bound | % Recovery |
|---|---|---|---|---|---|---|
| 100 | 85.5 | 76.3 | 1.10 | 1.4 | 98.6 | 90.4 |
| | 87.3 | 71.3 | 1.23 | 1.7 | 98.3 | 84.7 |
| | 84.1 | 65.3 | 1.19 | 1.8 | 98.2 | 77.7 |
| Mean ± SD | 85.6±1.6 | 71.0±5.5 | 1.17±0.07 | 1.7±0.2 | 98.3±0.2 | 84.3±6.4 |
| 250 | 196 | 164 | 2.49 | 1.5 | 98.5 | 87.3 |
| | 182 | 162 | 3.26 | 2.0 | 98.0 | 87.0 |
| | 193 | 105 | 3.19 | 3.0 | 97.0 | 56.7 |
| Mean± SD | 191±7[a] | 144±34 | 2.98±0.42 | 2.2±0.8 | 97.8±0.8 | 77.0±17.6 |
| 500 | 361 | 297[b] | 10.0[d] | NC | NC | NC |
| | 359 | 291 | 5.57 | 1.9 | 98.1 | 82.0 |
| | 365 | 280 | 5.49 | 2.0 | 98.0 | 78.8 |
| Mean ± SD | 362±3[a] | 285[c] | 5.53[c] | 1.9[c] | 98.1[c] | 80.4[c] |

NC = Not Calculated.
SD = Standard Deviation.
a. % difference from nominal value >15%: 23.8 (human 250 ng/mL), 27.6 (human 500 ng/mL).
b. Unreliable value since the correspondent dialysate was found to be protein contaminated. Value discarded.
c. n=2, SD not calculated.
d. Value excluded from mean due to protein contamination (protein concentration detected=1.04 mg/mL).

In human plasma, plasma-protein binding of plitidepsin was independent of drug concentration (100, 200 and 500 ng/ml), estimated at 98%.

**Table S3. Pharmacokinetic Parameters of Total Radioactivity in Blood and Plasma Following a Single Bolus IV Dose of [$^{14}$C]Plitidepsin to Male and Female Rat at 0.2 mg/kg.*(9)***

| Matrix | Gender | $AUC_{0-t}$ (h • ng-eq/g) |
|---|---|---|
| Plasma | Male | 59.3 |
| | Female | 52.0 |
| Lung | Male | 31600 |
| | Female | 28800 |

$AUC_{0-t}$ = area under the plasma concentration-time curve (AUC) from the start of dosing (0) to the last quantifiable time point (t), which was always 72 h apart from female plasma where the last quantifiable timepoint occurred at 48 h post-dose .

**Table S4. Summary of Protocol-Specified Efficacy Endpoints.**

| Endpoint | Dose Cohort | | | |
|---|---|---|---|---|
| | **1.5 mg N=14**[A] | **2.0 mg N=15** | **2.5 mg N=15** | **Total N=44** |
| Patients discharged from hospital | n (%) | | | |
| Day 1 to Day 7 | 3 (21.4) | 2 (13.3) | 5 (33.3) | 10 (22.7) |
| Day 1 to Day 8 | 6 (42.9) | 9 (60.0) | 10 (66.7) | 25 (56.8) |
| Day 1 to Day 15 | 11 (78.6) | 14 (93.3) | 11 (73.3) | 36 (81.8) |
| Day 1 to Day 31 | 13 (92.9) | 14 (93.3) | 13 (86.7) | 40 (90.9) |
| Mortality from Day 1 to | | | | |
| Day 7 | --- | --- | --- | --- |
| Day 15 | --- | --- | --- | --- |
| Day 31 | 1 (7.1) | --- | 1 (6.7) | 2 (4.5) |
| Patients requiring invasive mechanical ventilation and/or ICU admission | | | | |
| Day 1 to Day 7 | 2 (14.3) | 1 (6.7) | 2 (13.3) | 5 (11.4) |
| Day 8 to Day 15 | 1 (7.1) | 1 (6.7) | 1 (6.7) | 3 (6.8) |
| Day 16 to Day 31 | 1 (7.1) | 1 (6.7) | 1 (6.7) | 3 (6.8) |
| Day 1 to Day 31 | 2 (14.3) | 1 (6.7) | 3 (20.0) | 6 (13.6) |
| Patients requiring non-invasive mechanical ventilation | | | | |
| Day 1 to Day 7 | 4 (28.6) | 0 | 1 (6.7) | 5 (11.4) |
| Day 8 to Day 15 | 3 (21.4) | 0 | 2 (13.3) | 5 (11.4) |
| Day 16 to Day 31 | 1 (7.1) | 1 (6.7) | 1 (6.7) | 3 (6.8) |
| Day 1 to Day 31 | 5 (35.7) | 1 (6.7) | 2 (13.3) | 8 (18.2) |
| Patients requiring oxygen therapy at | | | | |
| Day 7 | 12 (85.7) | 12 (80.0) | 11 (73.3) | 35 (79.5) |
| Day 15 | 4 (28.6) | 1 (6.7) | 4 (26.7) | 9 (20.5) |
| Day 31 | 0 | 2 (13.3) | 1 (6.7) | 3 (6.8) |
| Day 1 to Day 31 | 12 (85.7) | 12 (80.0) | 11 (73.3) | 35 (79.5) |
| Mean change in viral load from baseline to[B] | $\log_{10}$ copies/mL | | | |
| Day 4 | -1.46 | -1.92 | -1.62 | -1.67 |
| Day 7 | -3.13 | -2.69 | -2.71 | -2.84 |
| Day 15 | -5.47 | -3.62 | -3.72 | -4.24 |
| Day 31 | -6.06 | -4.71 | -4.45 | -5.00 |
| Mean time from baseline until undetectable viral load[B] | Days | | | |
| | 11 | 14 | 14 | 13 |

A: Patient who experienced an anaphylactic reaction during the first plitidepsin infusion had treatment discontinued and was not considered evaluable for efficacy

B: Results based on 42 patients at Day 4 (13 at 1.5 mg, 14 at 2.0mg, 15 at 2.5mg), 40 patients at Day 7 (13 at 1.5mg, 14 at 2.0 mg, 13 at 2.5 mg), 38 patients at Day 15 (12 at 1.5mg, 13 at 2.0mg, 13 at 2.5mg), and 39 patients at Day 31 (11 at 1.5mg, 14 at 2.0mg, 14 at 2.5mg)

**Table S5. Six-point Ordinal Scale Assessment, per dose cohort and pre-specified time-points.**

| | 1.5 mg | | 2 mg | | 2.5 mg | | Total | |
|---|---|---|---|---|---|---|---|---|
| | N | % | N | % | N | % | N | % |
| **Day 1** | | | | | | | | |
| 2 Hospital admission, not requiring supplemental oxygen | 6 | 42.9 | 6 | 40.0 | 4 | 26.7 | 16 | 36.4 |
| 3 Hospital admission, requiring supplemental oxygen | 8 | 57.1 | 9 | 60.0 | 11 | 73.3 | 28 | 63.6 |
| **Day 4** | | | | | | | | |
| 2 Hospital admission, not requiring supplemental oxygen | 4 | 28.6 | 6 | 40.0 | 5 | 33.3 | 15 | 34.1 |
| 3 Hospital admission, requiring supplemental oxygen | 8 | 57.1 | 9 | 60.0 | 10 | 66.7 | 27 | 61.4 |
| 4 Hospital admission, requiring high-flow nasal cannula or non-invasive mechanical ventilation | 2 | 14.3 | . | . | . | . | 2 | 4.5 |
| **Day 7** | | | | | | | | |
| 1 Discharged (Alive) | 3 | 21.4 | 2 | 13.3 | 5 | 33.3 | 10 | 22.7 |
| 2 Hospital admission, not requiring supplemental oxygen | 3 | 21.4 | 3 | 20.0 | 3 | 20.0 | 9 | 20.5 |
| 3 Hospital admission, requiring supplemental oxygen | 5 | 35.7 | 10 | 66.7 | 4 | 26.7 | 19 | 43.2 |
| 4 Hospital admission, requiring high-flow nasal cannula or non-invasive mechanical ventilation | 3 | 21.4 | . | . | 3 | 20.0 | 6 | 13.6 |
| **Day 15** | | | | | | | | |
| 1 Discharged (Alive) | 11 | 78.6 | 14 | 93.3 | 11 | 73.3 | 36 | 81.8 |
| 3 Hospital admission, requiring supplemental oxygen | 1 | 7.1 | . | . | 1 | 6.7 | 2 | 4.5 |
| 4 Hospital admission, requiring high-flow nasal cannula or non-invasive mechanical ventilation | 1 | 7.1 | . | . | 3 | 20.0 | 4 | 9.1 |
| 5 Hospital admission, requiring extracorporeal membrane oxygenation or invasive mechanical ventilation | 1 | 7.1 | 1 | 6.7 | . | . | 2 | 4.5 |
| **Day 31** | | | | | | | | |
| 1 Discharged (alive) | 12 | 85.7 | 14 | 93.3 | 12 | 80.0 | 38 | 86.4 |
| 2 Hospital admission, not requiring supplemental oxygen | 1 | 7.1 | . | . | 1 | 6.7 | 2 | 4.5 |
| 4 Hospital admission, requiring high-flow nasal cannula or non-invasive mechanical ventilation | . | . | 1 | 6.7 | . | . | 1 | 2.3 |
| 5 Hospital admission, requiring extracorporeal membrane oxygenation or invasive mechanical ventilation | . | . | . | . | 1 | 6.7 | 1 | 2.3 |
| 6 Death | 1 | 7.1 | . | . | 1 | 6.7 | 2 | 4.5 |

**Table S6. Summary of Protocol-Specified Efficacy Endpoints, by disease severity at baseline.**

| Endpoint from day 1 to | | Mild | | Moderate | | Severe | |
|---|---|---|---|---|---|---|---|
| | | N | % | N | % | N | % |
| Patients discharged from hospital by, n (%) | Day 7 | 1 | 16.7 | 9 | 39.1 | . | . |
| | Day 8 | 4 | 66.7 | 17 | 73.9 | 4 | 26.7 |
| | Day 15 | 6 | 100.0 | 22 | 95.7 | 8 | 53.3 |
| | Day 31 | 6 | 100.0 | 23 | 100.0 | 11 | 73.3 |
| Patients requiring invasive mechanical ventilation and/or ICU admission at, n (%) | Day 7 | . | . | . | . | 5 | 33.3 |
| | Day 15 | . | . | . | . | 6 | 40.0 |
| | Day 31 | . | . | . | . | 6 | 40.0 |
| Patients requiring non-invasive mechanical ventilation at, n (%) | Day 7 | . | . | 1 | 4.3 | 4 | 26.7 |
| | Day 15 | . | . | 1 | 4.3 | 6 | 40.0 |
| | Day 31 | . | . | 1 | 4.3 | 7 | 46.7 |
| Patients requiring ICU admission at, n (%) | Day 7 | . | . | . | . | 5 | 33.3 |
| | Day 15 | . | . | . | . | 6 | 40.0 |
| | Day 31 | . | . | . | . | 6 | 40.0 |
| Patients requiring oxygen therapy at, n (%) | Day 7 | 2 | 33.3 | 18 | 78.3 | 15 | 100.0 |
| | Day 15 | 2 | 33.3 | 18 | 78.3 | 15 | 100.0 |
| | Day 31 | 2 | 33.3 | 18 | 78.3 | 15 | 100.0 |
| Patients requiring oxygen therapy (high flow) at, n(%) | Day 7 | . | . | 1 | 4.3 | 4 | 26.7 |
| | Day 15 | . | . | 1 | 4.3 | 6 | 40.0 |
| | Day 31 | . | . | 1 | 4.3 | 6 | 40.0 |
| Patients requiring oxygen therapy (low flow) at, n(%) | Day 7 | 2 | 33.3 | 18 | 78.3 | 15 | 100.0 |
| | Day 15 | 2 | 33.3 | 18 | 78.3 | 15 | 100.0 |
| | Day 31 | 2 | 33.3 | 18 | 78.3 | 15 | 100.0 |
| Patients requiring invasive mechanical ventilation at, n (%) | Day 15 | . | . | . | . | 2 | 13.3 |
| | Day 31 | . | . | . | . | 3 | 20.0 |
| Deaths at, n (%) | Day 31 | . | . | . | . | 2 | 13.3 |

**Table S7. Individual Viral Load Assessment at Pre-specified Timepoints.**

| Dose (mg/d) | Day of assessment | Result | $\log_{10}$ viral load copies/ mL |
|---|---|---|---|
| 1.5 | Baseline | Not detected | 0.00 |
| | Day 4 | | . |
| | Day 7 | | . |
| | Day 15 | | . |
| | Day 31 | | . |
| 1.5 | Baseline | Positive | 8.36 |
| | Day 4 | Positive | 5.14 |
| | Day 7 | Positive | 8.47 |
| | Day 15 | Not detected | 0.00 |
| | Day 31 | Positive | 4.42 |
| 1.5 | Baseline | Positive | 9.70 |
| | Day 4 | Positive | 9.54 |
| | Day 7 | Positive | 4.70 |
| | Day 15 | Not detected | 0.00 |
| | Day 31 | Not detected | 0.00 |
| 2 | Baseline | Positive | 7.03 |
| | Day 4 | Positive | 6.17 |
| | Day 7 | Positive | 4.02 |
| | Day 15 | Not detected | 0.00 |
| | Day 31 | Not detected | 0.00 |
| 1.5 | Baseline | Positive | 3.19 |
| | Day 4 | Not detected | 0.00 |
| | Day 7 | Not detected | 0.00 |
| | Day 15 | Not detected | 0.00 |
| | Day 31 | Not detected | 0.00 |
| 2 | Baseline | Positive | 5.12 |
| | Day 4 | Positive | 4.53 |
| | Day 7 | Not detected | 0.00 |
| | Day 15 | Not detected | 0.00 |
| | Day 31 | Not detected | 0.00 |
| 1.5 | Baseline | Positive | 4.92 |
| | Day 4 | Positive | 4.02 |

| Dose (mg/d) | Day of assessment | Result | $log_{10}$ viral load copies/ mL |
|---|---|---|---|
| | Day 7 | Not detected | 0.00 |
| | Day 15 | Not detected | 0.00 |
| | Day 31 | Not detected | 0.00 |
| 2 | Baseline | Positive | 6.50 |
| | Day 4 | Positive | 4.47 |
| | Day 7 | Positive | 3.62 |
| | Day 15 | Positive | 5.52 |
| | Day 31 | Positive | 4.31 |
| 1.5 | Baseline | Positive | 6.51 |
| | Day 4 | Positive | 6.56 |
| | Day 7 | Positive | 4.71 |
| | Day 15 | Not detected | 0.00 |
| | Day 31 | Not detected | 0.00 |
| 2.5 | Baseline | Positive | 10.09 |
| | Day 4 | Positive | 8.67 |
| | Day 7 | Positive | 7.29 |
| | Day 15 | Positive | 5.92 |
| | Day 31 | Not detected | 0.00 |
| 2 | Baseline | Positive | 5.66 |
| | Day 4 | Positive | 5.34 |
| | Day 7 | Positive | 4.33 |
| | Day 15 | Positive | 5.22 |
| | Day 31 | Positive | 4.69 |
| 2 | Baseline | Positive | 6.49 |
| | Day 4 | Positive | 5.99 |
| | Day 7 | Positive | 2.89 |
| | Day 15 | Not detected | 0.00 |
| | Day 31 | Not detected | 0.00 |
| 1.5 | Baseline | Not detected | 0.00 |
| | Day 4 | Not detected | 0.00 |
| | Day 7 | Not detected | 0.00 |
| 2.5 | Baseline | Positive | 10.62 |

| **Dose (mg/d)** | **Day of assessment** | **Result** | **$log_{10}$ viral load copies/ mL** |
|---|---|---|---|
| | Day 4 | Positive | 9.84 |
| | Day 7 | Positive | 9.54 |
| | Day 15 | Positive | 8.33 |
| | Day 31 | | . |
| 2 | Baseline | Positive | 6.96 |
| | Day 4 | Positive | 5.69 |
| | Day 7 | Positive | 5.20 |
| | Day 15 | Positive | 4.43 |
| | Day 31 | Not detected | 0.00 |
| 1.5 | Baseline | Positive | 7.20 |
| | Day 4 | Positive | 5.76 |
| | Day 7 | Positive | 3.82 |
| | Day 15 | Not detected | 0.00 |
| | Day 31 | Not detected | 0.00 |
| 2.5 | Baseline | Positive | 5.18 |
| | Day 4 | Not detected | 0.00 |
| | Day 7 | Not detected | 0.00 |
| | Day 15 | Not detected | 0.00 |
| | Day 31 | Not detected | 0.00 |
| 2 | Baseline | Positive | 6.20 |
| | Day 4 | Positive | 5.50 |
| | Day 7 | Positive | 5.49 |
| | Day 15 | Positive | 4.92 |
| | Day 31 | Not detected | 0.00 |
| 2.5 | Baseline | Not detected | 0.00 |
| | Day 4 | Positive | 3.79 |
| | Day 7 | Not detected | 0.00 |
| | Day 15 | Not detected | 0.00 |
| | Day 31 | Not detected | 0.00 |
| 2.5 | Baseline | Positive | 8.76 |
| | Day 4 | Positive | 9.25 |
| | Day 7 | Positive | 6.99 |
| | Day 15 | | . |

| Dose (mg/d) | Day of assessment | Result | $log_{10}$ viral load copies/ mL |
|---|---|---|---|
| | Day 31 | Not detected | 0.00 |
| 2 | Baseline | Positive | 7.01 |
| | Day 4 | Positive | 7.95 |
| | Day 7 | Positive | 6.52 |
| | Day 15 | | . |
| | Day 31 | Not detected | 0.00 |
| 2.5 | Baseline | Positive | 6.39 |
| | Day 4 | Positive | 4.30 |
| | Day 7 | Not detected | 0.00 |
| | Day 15 | Not detected | 0.00 |
| | Day 31 | Not detected | 0.00 |
| 2 | Baseline | | . |
| | Day 4 | Not detected | 0.00 |
| | Day 7 | Not detected | 0.00 |
| | Day 15 | Not detected | 0.00 |
| | Day 31 | Not detected | 0.00 |
| 2 | Baseline | Positive | 3.80 |
| | Day 4 | Not detected | 0.00 |
| | Day 7 | Positive | 4.68 |
| | Day 15 | Not detected | 0.00 |
| | Day 31 | Not detected | 0.00 |
| 2.5 | Baseline | Not detected | 0.00 |
| | Day 4 | Not detected | 0.00 |
| | Day 7 | Not detected | 0.00 |
| | Day 15 | Not detected | 0.00 |
| | Day 31 | Not detected | 0.00 |
| 2.5 | Baseline | Positive | 5.70 |
| | Day 4 | Not detected | 0.00 |
| | Day 7 | | . |
| | Day 15 | | . |
| | Day 31 | Positive | 3.60 |
| 2.5 | Baseline | Positive | 4.64 |
| | Day 4 | Positive | 5.45 |

| **Dose (mg/d)** | **Day of assessment** | **Result** | **$\log_{10}$ viral load copies/ mL** |
|---|---|---|---|
| | Day 7 | Positive | 3.05 |
| | Day 15 | Not detected | 0.00 |
| | Day 31 | Not detected | 0.00 |
| 1.5 | Baseline | Positive | 5.77 |
| | Day 4 | Positive | 3.25 |
| | Day 7 | Not detected | 0.00 |
| | Day 15 | Positive | 6.05 |
| | Day 31 | Not detected | 0.00 |
| 2.5 | Baseline | Positive | 6.72 |
| | Day 4 | Positive | 4.37 |
| | Day 7 | Not detected | 0.00 |
| | Day 15 | Not detected | 0.00 |
| | Day 31 | Not detected | 0.00 |
| 2 | Baseline | Positive | 6.16 |
| | Day 4 | Not detected | 0.00 |
| | Day 7 | Not detected | 0.00 |
| | Day 15 | Not detected | 0.00 |
| | Day 31 | Not detected | 0.00 |
| 1.5 | Baseline | Positive | 7.76 |
| | Day 4 | Positive | 8.40 |
| | Day 7 | Positive | 6.63 |
| | Day 15 | Not detected | 0.00 |
| | Day 31 | Not detected | 0.00 |
| 2.5 | Baseline | Positive | 6.00 |
| | Day 4 | Positive | 4.77 |
| | Day 7 | Positive | 4.44 |
| | Day 15 | Not detected | 0.00 |
| | Day 31 | Not detected | 0.00 |
| 1.5 | Baseline | Positive | 6.90 |
| | Day 4 | Positive | 6.54 |
| | Day 7 | Positive | 4.34 |
| | Day 15 | Not detected | 0.00 |
| | Day 31 | Not detected | 0.00 |

| Dose (mg/d) | Day of assessment | Result | $log_{10}$ viral load copies/ mL |
|---|---|---|---|
| 2 | Baseline | Positive | 5.76 |
| | Day 4 | Positive | 4.46 |
| | Day 7 | Not detected | 0.00 |
| | Day 15 | Positive | 8.14 |
| | Day 31 | Positive | 7.37 |
| 2.5 | Baseline | Positive | 8.04 |
| | Day 4 | Positive | 5.57 |
| | Day 7 | Positive | 5.49 |
| | Day 15 | Positive | 3.80 |
| | Day 31 | Not detected | 0.00 |
| 2 | Baseline | Positive | 4.70 |
| | Day 4 | Not detected | 0.00 |
| | Day 7 | Positive | 4.59 |
| | Day 15 | Not detected | 0.00 |
| | Day 31 | Not detected | 0.00 |
| 2.5 | Baseline | Positive | 5.16 |
| | Day 4 | Not detected | 0.00 |
| | Day 7 | Positive | 3.33 |
| | Day 15 | Not detected | 0.00 |
| | Day 31 | Not detected | 0.00 |
| 2.5 | Baseline | Positive | 5.45 |
| | Day 4 | Positive | 6.26 |
| | Day 7 | | . |
| | Day 15 | Positive | 5.66 |
| | Day 31 | Positive | 5.33 |
| 1.5 | Baseline | Positive | 4.56 |
| | Day 4 | Not detected | 0.00 |
| | Day 7 | Not detected | 0.00 |
| | Day 15 | Not detected | 0.00 |
| | Day 31 | Not detected | 0.00 |
| 1.5 | Baseline | Positive | 6.26 |
| | Day 4 | Positive | 3.61 |

| Dose (mg/d) | Day of assessment | Result | $log_{10}$ viral load copies/ mL |
|---|---|---|---|
| | Day 7 | Not detected | 0.00 |
| | Day 15 | Not detected | 0.00 |
| | Day 31 | Not detected | 0.00 |
| 2 | Baseline | Positive | 6.53 |
| | Day 4 | Positive | 5.30 |
| | Day 7 | Positive | 3.38 |
| | Day 15 | Not detected | 0.00 |
| | Day 31 | Not detected | 0.00 |
| 2.5 | Baseline | Positive | 3.82 |
| | Day 4 | Not detected | 0.00 |
| | Day 7 | Not detected | 0.00 |
| | Day 15 | Not detected | 0.00 |
| | Day 31 | Positive | 4.68 |
| 1.5 | Baseline | Positive | 8.05 |
| | Day 4 | Positive | 7.39 |
| | Day 7 | Positive | 5.83 |
| | Day 15 | Positive | 7.46 |
| | Day 31 | | . |
| 2 | Baseline | Positive | 4.39 |
| | Day 4 | Not detected | 0.00 |
| | Day 7 | Not detected | 0.00 |
| | Day 15 | Not detected | 0.00 |
| | Day 31 | Not detected | 0.00 |

**References and notes (Supplement)**


1. Fett C, DeDiego ML, Regla-Nava JA, Enjuanes L, Perlman S. Complete protection against severe acute respiratory syndrome coronavirus-mediated lethal respiratory disease in aged mice by immunization with a mouse-adapted virus lacking E protein. J Virol. 2013;87(12):6551-9.
2. DeDiego ML, Alvarez E, Almazan F, Rejas MT, Lamirande E, Roberts A, et al. A severe acute respiratory syndrome coronavirus that lacks the E gene is attenuated in vitro and in vivo. J Virol. 2007;81:1701-13.
3. Livak KJ, Schmittgen TD. Analysis of relative gene expression data using real-time quantitative PCR and the 2(-Delta Delta C(T)) Method. Methods. 2001;25:402-8.
4. Rodon J, Muñoz-Basagoiti J, Perez-Zsolt D, Noguera-Julian M, Paredes R, Mateu L, et al. Identification of Plitidepsin as Potent Inhibitor of SARS-CoV-2-Induced Cytopathic Effect after a Drug Repurposing Screen. bioRxiv. 2021:2020.04.23.055756.
5. Thorne LG, Reuschl A-K, Zuliani-Alvarez L, Whelan MVX, Noursadeghi M, Jolly C, et al. SARS-CoV-2 sensing by RIG-I and MDA5 links epithelial infection to macrophage inflammation. bioRxiv. 2020:2020.12.23.424169.
6. Kim S, Lee J. Boryung [Data on File]. Evaluation of antiviral activity of plitidepsin (Aplidin) against SARS-CoV-2. Study IPKBR-20200512. 2020:14.
7. Goutelle S, Maurin M, Rougier F, Barbaut X, Bourguignon L, Ducher M, et al. The Hill equation: a review of its capabilities in pharmacological modelling. Fundamental & clinical pharmacology. 2008;22(6):633-48.
8. PharmaMar. [Data on file] - Plitidepsin: In Vitro Determination of Plasma Protein Binding in Rat, Dog and Human Using Equilibrium Dialysis. VPT2678/2014. 2014.
9. PharmaMar. [Data on file] - [$^{14}$C]Plitidepsin: Pharmacokinetics, Disposition and Tissue Distribution in the Rat Following Single Intravenous Administration. VPT1992/2014. 2014.
10. Baker EH, Gnjidic D, Kirkpatrick CMJ, Pirmohamed M, Wright DFB, Zecharia AY. A call for the appropriate application of clinical pharmacological principles in the search for safe and efficacious COVID-19 (SARS-COV-2) treatments. Br J Clin Pharmacol. 2020;87:707-11.
11. Nalda-Molina R, Valenzuela B, Ramon-Lopez A, Miguel-Lillo B, Soto-Matos A, Perez-Ruixo JJ. Population pharmacokinetics meta-analysis of plitidepsin (Aplidin) in cancer subjects. Cancer Chemother Pharmacol. 2009;64(1):97-108.
